# Efficacy and safety of PCSK9 inhibitors for children and adolescents with heterozygous familial hypercholesterolaemia: Systematic review and meta-analysis of randomised controlled trials

**DOI:** 10.64898/2026.08.04.26359681

**Authors:** Alexis Llewellyn, Mark Simmonds, David Marshall, Melissa Harden, Steve E Humphries, Beth Woods, Manuel Gomes, Lorraine Priestley-Barnham, Uma Ramaswami, Mark Fisher, Nadeem Qureshi, Laila J. Tata

**Affiliations:** Centre for Reviews and Dissemination, University of York, UK; Institute of Cardiovascular Science, Faculty of Population Health, University College London, UK; Centre for Health Economics, University of York, UK; Department of Primary Care and Population Health, University College London, UK; Harefield Hospital, Guy’s and St Thomas NHS Trust, UK; Royal Free Hospital and Genetics and Genomic Medicine, University College London, UK; Independent, UK; Centre for Academic Primary Care, University of Nottingham, UK and NIHR School of Primary Care Research, University of Nottingham, UK; Lifespan and Population Health Unit and Centre for Perinatal Research, School of Medicine, University of Nottingham, UK

## Abstract

**Background:** Statins and ezetimibe are the preferred lipid-lowering therapies (LLTs) for children with heterozygous familial hypercholesterolaemia (HeFH). Proprotein convertase subtilisin/kexin type 9 inhibitors (PCSK9i) are newer add-on therapies for individuals not achieving low-density lipoprotein-cholesterol (LDL-C) targets. We evaluated the efficacy and safety of PCSK9i in children aged <18 years with HeFH.

**Methods:** Systematic review and pairwise meta-analyses of randomised-controlled trials (RCTs) of evolocumab, alirocumab and inclisiran. Comprehensive bibliographic searches were conducted in February 2026. Risk of bias was assessed with Cochrane RoB 2.

**Results:** Of 2798 unique records screened, three RCTs were included (n=451, mean age 13 years, follow-up 24 to 47 weeks). Each trial evaluated either evolocumab, alirocumab or inclisiran against placebo as add-on to baseline LLT. Participants had elevated LDL-C (>3.4 mmol/L [130 mg/dL]) despite stable LLT. Overall risk of bias was low. PCSK9i reduced LDL-C by an average of 35.44% (95% CI −41.74 to −29.14, I^2^=50.8%) and by 1.63 mmol/L [62.93 mg/dL] (95% CI −1.86 to −1.39, I^2^=18.6%) compared with placebo. There was no evidence of differences between PCSK9i and placebo in tolerability, growth and maturation, and overall incidence of adverse events.

**Conclusions:** PCSK9i add-on therapy leads to substantial reductions in LDL-C in paediatric patients with HeFH failing to achieve LDL-C targets with standard LLT. While the findings of this review support the use of PCSK9i in a subset of children and young people with HeFH, limited trial numbers and short follow-up periods underscore the need for future high-quality studies evaluating long-term safety, effectiveness and cost-effectiveness.

GRAPHICAL ABSTRACT
PCSK9 inhibitors for children and adolescents with HeFH

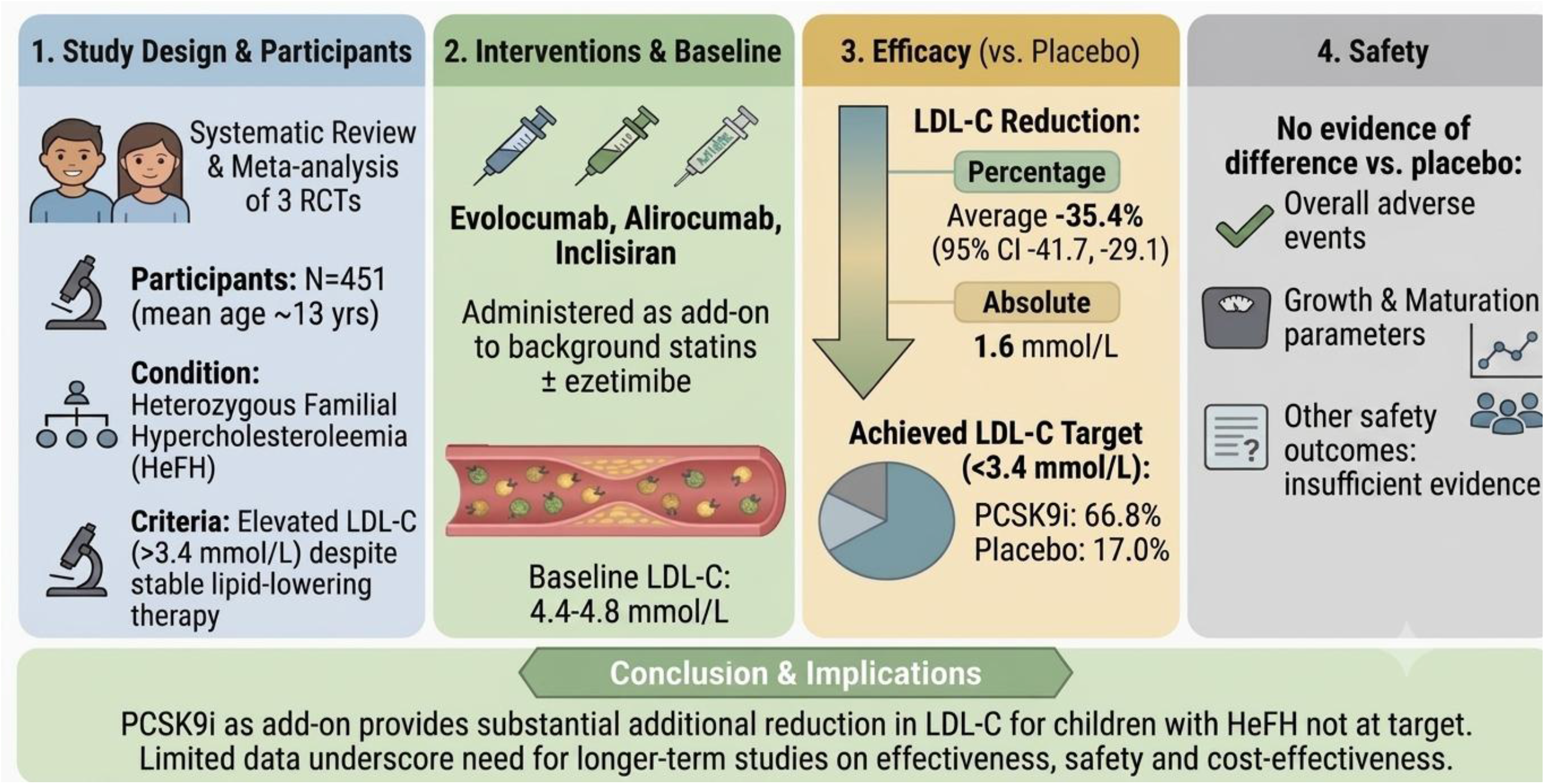

## Background

Familial hypercholesterolaemia (FH) is one of the most common inherited metabolic disorders. Heterozygous familial hypercholesterolaemia (HeFH), an autosomal dominant condition characterised by lifelong elevation of low-density lipoprotein cholesterol (LDL-C), affects approximately 1 in 300 children and adults worldwide. ^1, 2^ Patients with HeFH typically have LDL-C concentrations two-to three-fold higher than those observed in the general population, resulting in accelerated atherosclerosis and an increased risk of premature cardiovascular disease (CVD). Subclinical vascular changes may be detectable from childhood, and cumulative exposure to elevated LDL-C (“cholesterol burden”) is a major determinant of future atherosclerotic cardiovascular disease (ASCVD) risk. ^3–5^

Timely initiation of lipid-lowering therapy (LLT) is recommended to reduce lifelong LDL-C exposure and future cardiovascular risk. Statins remain first-line pharmacological therapy for children and adolescents with HeFH. The 2026 European Atherosclerosis Society consensus statement recommends starting LLT from ideally 6 years of age, to reduce LDL to ≤3.5 mmol/L (135mg/dL) before 10 years, and to ≤3.0 mmol/L (116 mg/dL) at 10 to 18 years or before age 10 for children with major risk factors.^6^ US guidelines support the use of statins from 8 years of age if lifestyle management is insufficient to reduce LDL-C below 4.1 mmol/L (160 mg/dL); statins and other LLTs may be considered at 6 years or at lower LDL-C thresholds for children with additional clinical or family risk factors. A reduction of ≥50% in LDL-C from baseline and a target LDL-C of ≤3.4 mmol/L (130 mg/dL) is generally advised, which can be lowered to ≤ 2.6 mmol/L (100 mg/dL) in children with additional risk factors. ^7^ Ezetimibe is recommended as adjunctive therapy when LDL-C targets are not achieved, or as an alternative in the relatively uncommon case of statin intolerance. ^8 6 9^

Despite optimised LLT on statins and/or ezetimibe, a proportion of paediatric patients with HeFH remain above recommended LDL-C thresholds, highlighting the need for additional LLTs.^6, 8^ Proprotein convertase subtilisin/kexin type 9 inhibitors (PCSK9i) enhance hepatic LDL receptor recycling and LDL-C clearance. Monoclonal antibodies targeting circulating PCSK9, including evolocumab and alirocumab, have shown additional reductions in LDL-C concentrations in adults and paediatric HeFH populations receiving background LLT.^10 11 12, 13^ Inclisiran, a small interfering RNA therapy that suppresses hepatic PCSK9 synthesis, has also recently been evaluated in adolescents with HeFH.^14^ The EAS recommends that PCSK9i are considered instead of ezetimibe if LDL-C concentrations are not close to goal [i.e. ≥4.0 mmol/L (155 mg/dL)] on statin monotherapy, with the intention to prescribe a maximum of two drugs rather than three drugs where possible.^6^ Although valuable, previous systematic reviews of PCSK9i in paediatric populations did not include trial evidence for inclisiran,^15–17^ and in some instances pooled heterogeneous populations with potentially distinct risk profiles, co-interventions and responses to therapy (such as HoFH and HeFH, or adults and children).^15, 17^ Whilst systematic reviews have shown good efficacy and safety in children with FH, the current systematic review evidence on LLTs for children with HeFH is not up to date. This systematic review aims to evaluate all available evidence on the efficacy and safety of PCSK9-targeted therapies in children and adolescents with HeFH.

## Methods

This systematic review and meta-analysis was undertaken in accordance with the Centre for Reviews and Dissemination guidance on systematic reviews and reported following PRISMA guidance.^18, 19^ The review was registered prospectively in PROSPERO and conducted as part of a wider review of LLT for paediatric HeFH (CRD420251063400).^20^ Although the wider review searched for trials of statins, ezetimibe and PCSK9i, no new studies of statins and ezetimibe were identified since our initial published systematic review, which focused exclusively on statins and ezetimibe (CRD42023408037).^20 21^ Following a feasibility assessment, it was considered that the trial evidence for PCSK9i was too heterogeneous to be pooled with existing trials of statins and ezetimibe. In particular, trials of statins and ezetimibe included patients on first-line therapy with substantially higher baseline LDL-C, whereas all existing trials of PCSK9i evaluated this therapy as add-on to statins/ezetimibe in children who had failed to reach recommended LDL-C targets despite stable statins/ezetimibe therapy. Therefore, this systematic review report focuses solely on PCSK9i trials.

### Search strategy and study selection

Literature searches were conducted in February 2026 using MEDLINE (Ovid). Embase (Ovid), the Cochrane Library (Wiley), Science Citation Index (Web of Science), the INAHTA database. Additional searches included clinical trial registries, unpublished and ongoing studies, conference proceedings and citation screening of relevant reviews and included studies. The search strategy is presented in a Supplementary File.

Eligible studies were randomised controlled trials (RCTs) evaluating PCSK9 inhibitors in children and young people aged up to 18 years with clinically diagnosed HeFH according to genetic testing and/or serum lipid testing and family history. Studies of homozygous familial hypercholesterolaemia were excluded.

Outcomes of interest included LDL-C change from baseline (absolute or percentage), achievement of LDL-C targets, other lipid parameters (total cholesterol [TC], high-density lipoprotein cholesterol [HDL-C], carotid intima-media thickness (cIMT), growth and maturation measures, and adverse events. Two reviewers independently screened titles, abstracts, and full texts against predefined eligibility criteria, with disagreements resolved by discussion.

### Data extraction and quality assessment

All data on study characteristics, interventions, and outcomes were extracted by one reviewer and checked by a second reviewer from published records and trial protocols. Risk of bias was assessed using the Cochrane risk of bias (RoB 2) tool.^22^

### Data synthesis

Meta-analysis was undertaken where at least two studies contributed data for an outcome. Random-effects meta-analyses were performed with between-study variance estimated using the restricted maximum likelihood (REML) method. Fixed effect models were run as sensitivity analyses. Continuous outcomes were synthesised as mean differences and dichotomous outcomes as relative risks. Where reported, adjusted trial estimates (i.e. least-squares mean differences derived from mixed models for repeated measures) were preferentially pooled. Pooled analyses of absolute change in LDL-C were based on final values as specified in the protocol, to assess longest available follow-up within the randomised period. Statistical heterogeneity was evaluated using the I^2^ statistic, τ^2^ and inspection of between-study heterogeneity estimates, accounting for data sparsity. For studies reporting multiple follow-up assessments, data from the latest blinded follow-up were used as specified in the protocol. Results were presented in forest plots, with additional data presented in supplementary tables. Publication bias was not formally assessed due to the limited number of trials. Analyses were conducted in R using the meta package (version 8.5-0).

PCSK9i trial participants had prior exposure to LLT, and substantially lower baseline LDL-C concentrations compared with existing trials of statins/ezetimibe,^21^ and were therefore considered clinically distinct. Following assessment of network feasibility, it was considered that PCSK9i trials populations were too distinct from trials of statins/ezetimibe to be combined in the same network.

The evidence base for PCSK9i was considered too sparse to support a robust, separate network meta-analysis for this treatment class, and only pairwise meta-analyses were conducted. Where limited or clinically heterogeneous evidence precluded meta-analysis, findings were summarised narratively and in tables. Classifications of safety events according to seriousness or causality were based on the assessments of the investigators in the included studies.

## Results

The study selection process is reported in Supplementary Figure 1. Of 2798 unique records screened, 134 full text reports were assessed for eligibility. Three unique trials were included from four published reports.^12–14, 23^ One trial (Santos et al 2024)^13^ reported on two separate PCSK9i regimens with separate comparator arms (injections every two weeks [Q2W] or every four weeks [Q4W]), and these were treated as distinct comparisons in the meta-analyses. Open-label continuation phases of included trials were excluded as they assessed outcomes beyond the randomised period; they are discussed below (discussion section).^14, 26^ No new trials of statins or ezetimibe were identified since our initial systematic review.

Overall risk of bias was low. Results of the risk of bias assessment are reported in Supplementary File. Table 1 presents a summary of the design and participant characteristics of included studies. All three trials were multicentre, international, double-blind placebo controlled RCTs, published between 2020 and 2026. They included a total of 451 patients under the age of 18 years with a diagnosis of HeFH (genetic or clinical) who had failed to reach LDL-C target at baseline (>3.4 mmol/L [130 mg/dL]) despite receiving an optimised dose of statins and/or ezetimibe. Investigator definitions of optimal doses of standard of care (SoC) LLT and participant selection criteria are reported in Supplementary Table 1. Optimal SoC LLT was defined as maximally tolerated statin treatment with or without additional LLT (e.g. ezetimibe) according to local/regional guidelines for at least four weeks prior to screening and was required to remain stable throughout the randomised period in all trials. Between 92.9% and 99.4% participants were on statins at baseline, and between 13.4% and 23.7% received ezetimibe. Only one study^12^ reported on statins dose strength, with 16.3% receiving a high dose, 61% moderate and 21% a low or unknown dose, according to classification from American guidelines.^24^ Mean age was between 12.8 and 13.7 years, and mean baseline LDL-C ranged from 170 mg/dL to 185 mg/dL across trial arms.

**Table 1.**
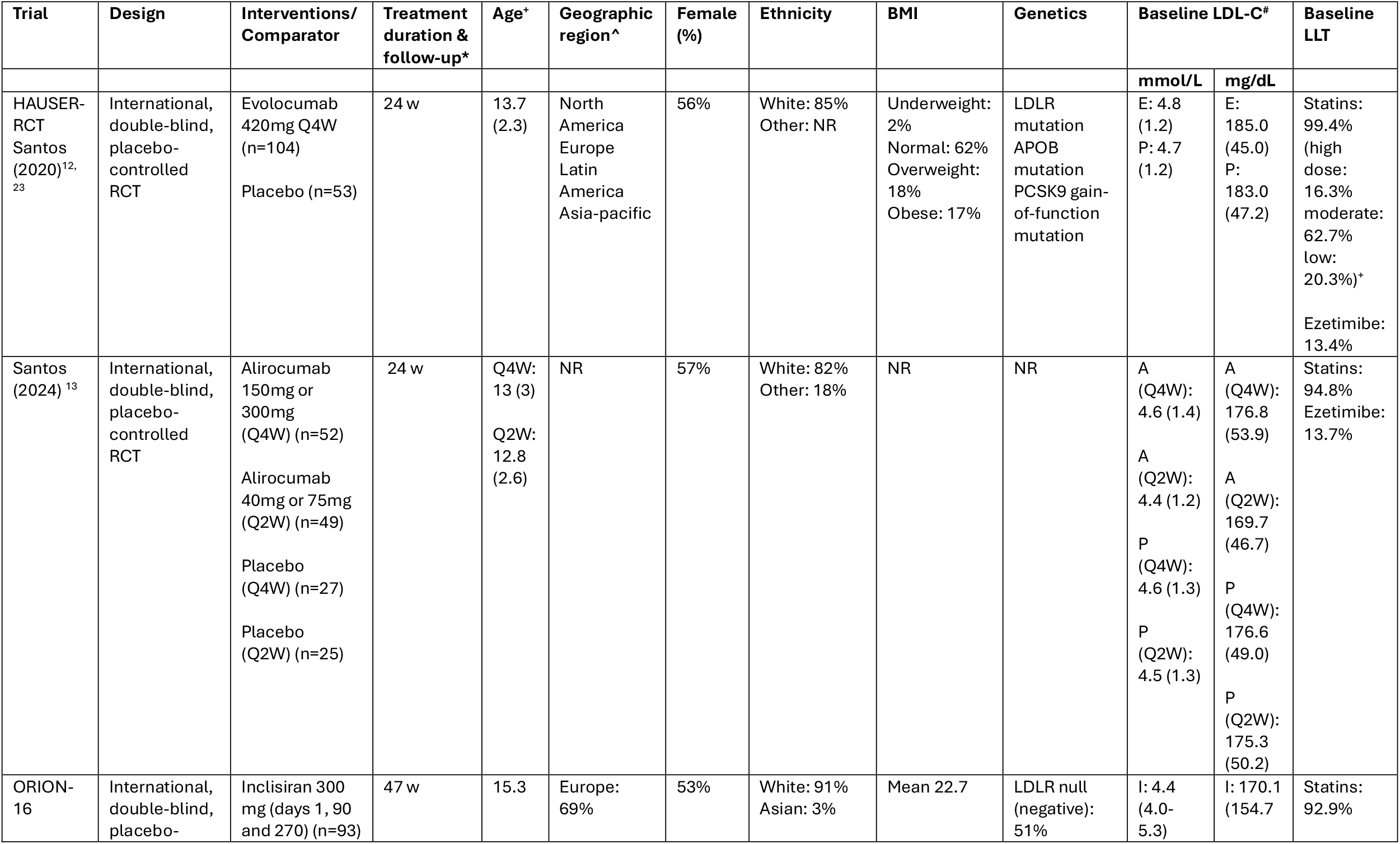
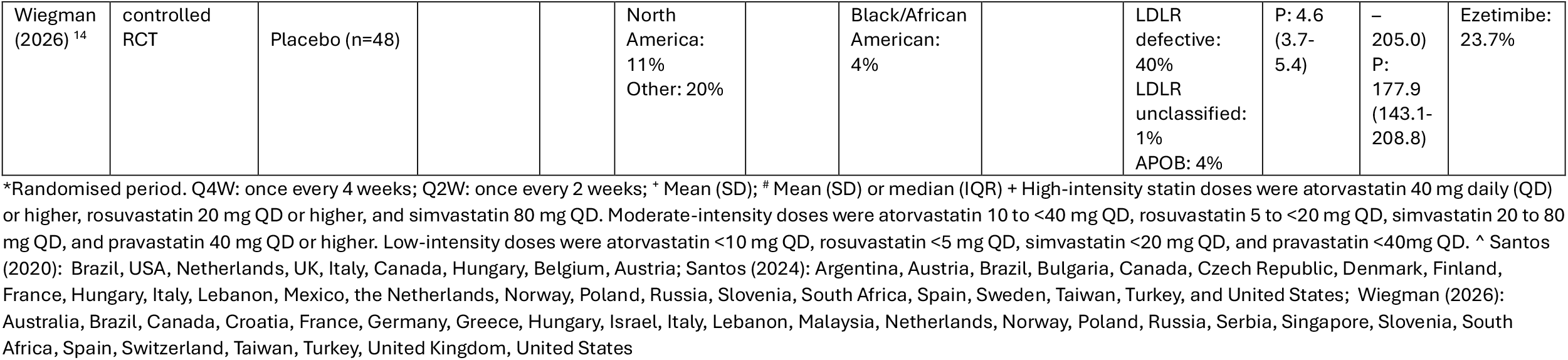
Characteristics of included studies.

### Clinical effectiveness

#### LDL-C

Figure 1 presents the results of the meta-analysis for least-square (LS) mean difference in percentage change from baseline between PCSK9i and placebo. PCSK9i reduced LDL-C by a mean of 35.44% more than placebo (95% CI −41.74 to −29.14, I^2^=50.8%)). Fixed effect sensitivity analyses yielded very similar results (LS mean difference −34.96, 95% CI −39.24 to −30.69).

**Figure 1.**
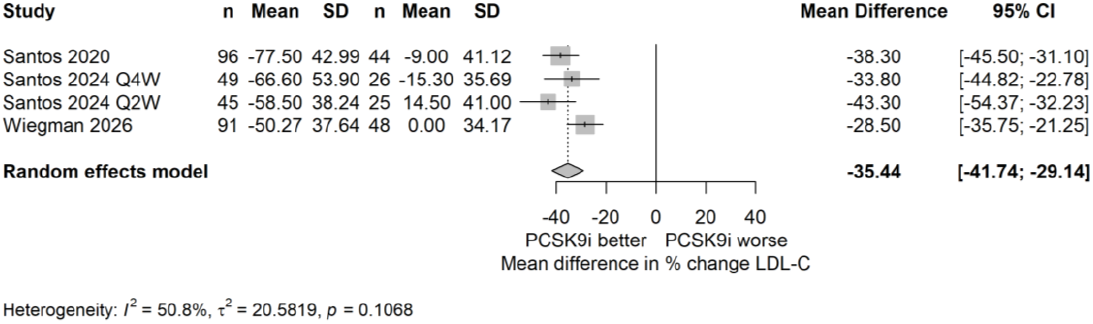
LS mean percentage change from baseline in LDL-C

Figure 2 shows PCSK9i reduced LDL-C by, on average, 1.56 mmol/L (95% CI −1.85 to −1.26, I^2^= 49.9%), or equivalently, 60.19 mg/dL (95% −71.69 to −48.69) more than placebo (Supplementary Figure 2). Further results are reported in Supplementary Tables 2 & 3. Sensitivity analyses yielded very similar results (−1.53 mmol/L, 95% CI −1.74 to −1.33, or −59.25 mg/dl, 95% CI −67.14 to −51.35).

**Figure 2.**
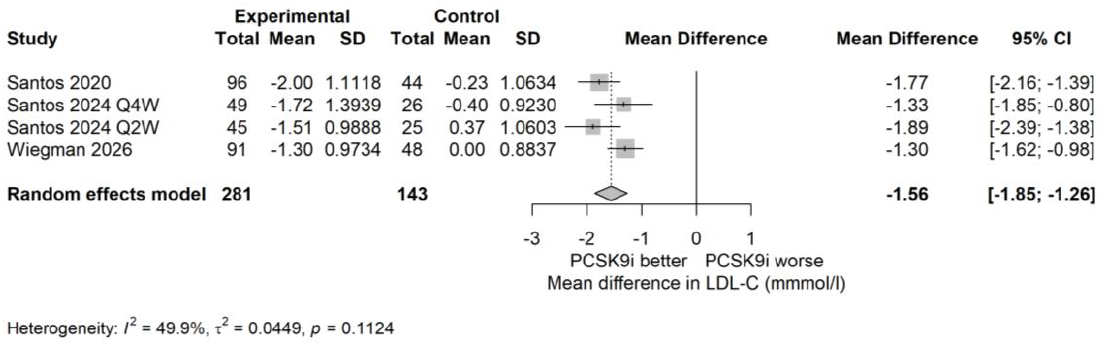
LS mean change in absolute LDL-C (mmol/L)

Figure 3 indicates that a total of 66.8% (199/298) of participants on PCSK9i reduced their LDL-C to below 130 mg/dL at follow-up, compared with 17.0% (26/153) in the placebo groups (RR 3.91, 95% CI 2.73 to 5.59, I^2^=0). Further LDL-C target results for individual studies are reported in Supplementary Table 4.

**Figure 3.**
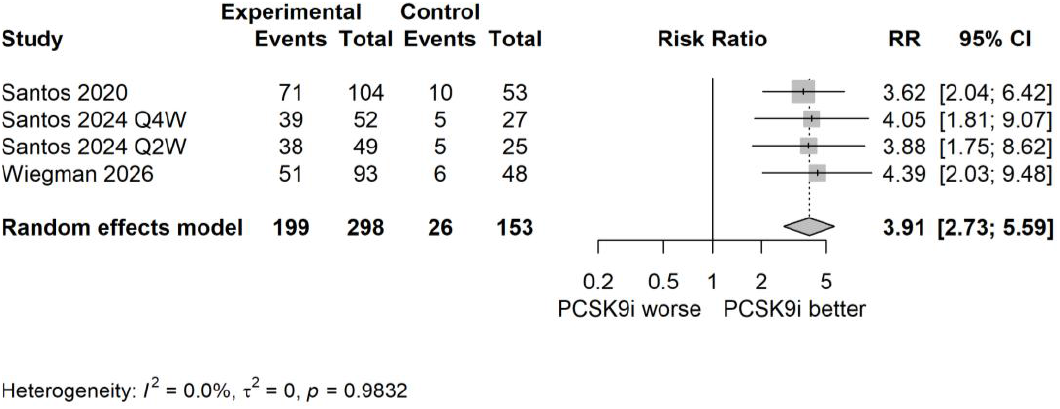
Reaching <130mg/dl at follow-up

#### Other cholesterol markers

PCSK9i reduced TC by 24.72% (95% CI −29.94 to −19.49, I^2^=55.5%) more than placebo (Supplementary Figure 3). One study reported numerically higher absolute TC reductions from baseline with inclisiran than placebo (inclisiran: −1.28 mmol/l [SD 1.23], or −49.4 mg/dl [SD 47.8], placebo: −0.02 mmol/l [SD 1.0] or −0.9mg/dl [SD 38.5], statistical significance not reported). No other studies reported absolute TC results therefore this outcome was not pooled.

Two studies reported on HDL-C at follow-up.^13 14^ Both reported an increase in HDL-C from baseline compared with placebo (Supplementary Table 5). Compared with placebo, the difference in HDL-C improvement was statistically significant for alirocumab Q2W, but not for alirocumab Q4W. Statistical significance was not reported for inclisiran vs. placebo. Two studies reported on triglycerides at follow-up.^13 14^ Variance estimates were wide and there was no evidence of a statistically significant difference between PCSK9i and placebo (Supplementary Table 6).

#### cIMT

One study measured cIMT and found no difference in changes from baseline between evolocumab and placebo at 24 weeks.^12^ No formal statistical comparisons were reported.

#### Growth and maturation

Meta-analyses of two studies showed no difference in weight and height changes for alirocumab and inclisiran compared with placebo at follow-up (Supplementary Figures 4 and 5).^13, 14^ All included trials measured pubertal development as Tanner stage separately for male and female participants and found similar changes between PCSK9i and placebo.

### Safety

Figure 4 shows no difference in incidence of adverse events between PCSK9i and placebo (RR 1.02; 95% CI 0.88 to 1.19, I^2^=0%). Supplementary Figure 6 shows the occurrence of serious adverse events was 10/298 (3.3%) across PCSK9i arms compared with 3/153 (2.0%) across placebo arms; the difference was not statistically significant (RR 1.52; 95% CI 0.46 to 5.04, I^2^=0%). Two serious adverse events of syncope (fainting) in two separate patients were reported in the alirocumab Q4W (2/52, 3.8%) and the study investigators described them as related to treatment. No treatment-related serious events were found in the other two included trials.

**Figure 4.**
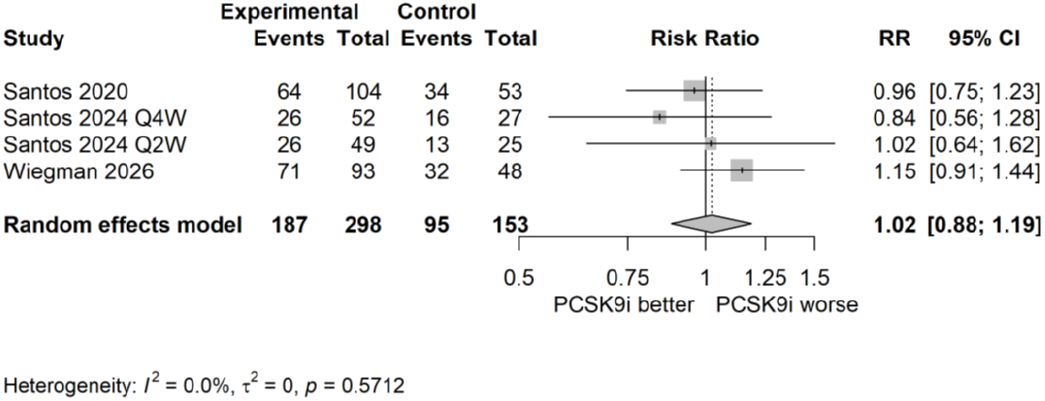
Adverse events (any)

Discontinuation due to adverse events occurred in 4/298 (1.3%) receiving PCSK9i compared to 0/153 placebo recipients. Reasons for discontinuation included syncope (n=1, alirocumab Q4W arm), arthropathy (n=1, non-serious, evolocumab arm), disturbance in attention and memory (n=1, non-serious, alirocumab Q4W arm), and vertigo and nausea (n=1, mild and transient, inclisiran arm).

Supplementary Figure 7 shows that the incidence of non-serious injection-related adverse events was higher across PCSK9i arms (21/298 [7.0%] vs. 3/153 [2.0%]) than placebo, although the difference was not statistically significant (RR 2.55; 95% CI 0.96 to 6.83, I^2^=0%). Sensitivity analyses yielded very similar results for all meta-analyses of safety outcomes. No antibodies against evolocumab/alirocumab were detected in the two trials of monoclonal antibodies.

Other adverse events of special interest included general allergic reaction (2/52 [3.8%], disturbance in attention and memory [1/52 [1.9%] and hypoesthesia [1/52 [1.9%] in the Q4W alirocumab arm (vs none in the alirocumab Q2W and Q4W/Q2W placebo arms). Changes from baseline in neurologic assessment with the Cogstate battery of cognitive tests at week 24 were similar in the evolocumab and placebo groups.

Supplementary Table 7 shows that no cases of elevated creatine kinase (>5x ULN), aminotransferases (>3 x ULN) or creatinine (>2 mg/dL) were found in either arm of the evolocumab trial. In the inclisiran trial, both treatment and placebo groups showed similarly low rates of creatine kinase (>5x ULN, 3% vs. 6% respectively) and total bilirubin (>2x ULN, 3% vs. 2%), with no elevations in creatinine (>2 mg/dL) or ALT (>3× ULN) in either group. Laboratory confirmed adverse events were not reported in the alirocumab trial.

## Discussion

This systematic review and meta-analysis included three randomised trials of PCSK9i in children with HeFH. All trials evaluated PCSK9i as add-on therapy to statins and/or ezetimibe standard of care therapy. Participants had persistent LDL-C elevations (>3.4 mmol/L [130 mg/dL]) at baseline despite stable, optimised standard of care lipid-lowering treatment.

Our meta-analyses showed a substantial benefit of PCSK9i inhibitors, with large reductions in serum LDL-C in both percentage and absolute terms. The two trials that reported on HDL-C also suggested a numerical increase in HDL-C compared with placebo, but this was inconclusive. Evidence was insufficient to draw conclusions on the benefits of PCSK9i on triglyceride levels and cIMT.

Approximately two-thirds of patients receiving PCSK9i reduced their LDL-C below 3.4 mmol/L (130 mg/dL), compared with 17% in the placebo group. Despite the use of PCSK9i as add-on to statins and/or ezetimibe, this means that a significant minority of patients still failed to reach the stricter EAS recommended target ≤3.0 mmol/L (115 mg/dL).^6^

LDL-C reductions in the paediatric patients were lower than those reported in adults. In adults with HeFH receiving maximally tolerated LLT, evolocumab (RUTHERFORD-2) reduced LDL-C by approximately 60%, alirocumab (ODYSSEY FH I/FH II) by 51–58%, and inclisiran (ORION-9) by about 47% compared with placebo.^10 11 25^ The reasons for these observed differences in response magnitude between adults and children are unclear, although naïve comparisons between age groups are limited by differences in study design, trial populations, dosage, and generally more conservative management in childcren and should therefore be interpreted with caution.

PCKS9i were generally well-tolerated and had no measurable effect on growth parameters, including weight, height and pubertal development at up to one year. Meta-analyses showed no evidence that PCSK9i led to a higher incidence of adverse events overall; there was no evidence of a statistically significant difference in serious adverse event rates between PCSK9i and placebo, although events were uncommon and meta-analyses were underpowered for this outcome. Two serious treatment-related adverse events occurred (syncope), both in the alirocumab Q4W arm, one of which led to treatment discontinuation. Open-label continuation studies have not identified additional safety concerns to date, although further safety monitoring is needed.^26 14^ Injection-site reactions were all reported as non-serious and were more frequent in PCSK9i recipients although the data were insufficient to accurately quantify the risk of this event.

### Strengths and limitations

To our knowledge, this is the first systematic review and meta-analysis of PCSK9i to include monoclonal antibodies as well as inclisiran in paediatric HeFH. Rigorous systematic review methods were applied following a pre-specified protocol and following established guidance. ^18–20^ Sensitivity analyses using fixed-effect models yielded results that were highly consistent with random-effects models, indicating that the findings were robust to model assumptions; there was no evidence of substantial heterogeneity. All included studies were at low overall risk of bias.

Our findings broadly align with previous systematic reviews in confirming that PCSK9-targeted therapies provide clinically meaningful reductions in LDL-C and have a favourable short-term safety profile in paediatric populations. This is the first systematic review and meta-analysis to incorporate newly available clinical trial evidence for the small interfering RNA therapy, inclisiran. While prior analyses frequently compromised clinical homogeneity by pooling distinct cohorts—such as mixing adult and pediatric data ^17^ or combining heterozygous (HeFH) and homozygous (HoFH) phenotypes, ^15, 17^ this review strictly isolates paediatric patients with HeFH, providing a more precise and targeted evaluation of efficacy and safety for this specific group.

Due to the limited number of studies, some pre-specified outcomes could not be pooled, our ability to assess and explore heterogeneity was limited (notably I^2^ values should be interpreted with caution)^27^ and analyses comparing the relative efficacy and safety of individual or class-specific (e.g. monoclonal antibodies and small interfering RNA) PCSK9i were not feasible. The age of participants and follow-up duration of paediatric trials meant that it was not possible to assess longer-term cardiovascular outcomes. The short follow-up duration and lack of data limited the ability to evaluate the effect of treatment on cIMT and longer-term safety outcomes.

While the included trials represent international, multicentre cohorts, the baseline demographics reveal a notable lack of ethnic diversity, with white participants comprising most (82% to 91%) of the pooled population. Reporting on geographic and socioeconomic metrics was inconsistent across trials, and specific genetic variations (such as *LDLR* null vs. defective mutations) were not uniformly reported.

Although all patients were reportedly receiving an optimal dose of statins, only one study reported baseline statins dose strength explicitly.^12^ In HAUSER-RCT, use of high-intensity statins (16.3%) and ezetimibe (13.4%) were relatively infrequent, and ezetimibe use was similarly infrequent in the other included trials.^13 14 28^ Current lipid-management guidelines recommend maximally tolerated statin therapy as first-line treatment for severe LDL-C elevation, with ezetimibe generally recommended before escalation to PCSK9i therapy.[EAS 2026] ^8^ Consequently, it is possible that some participants had not received fully optimised conventional lipid-lowering therapy before trial enrolment. However, this may reflect routine care, where many children with HeFH do not receive intensive lipid-lowering regimens despite evidence supporting the short-term safety and tolerability of higher-intensity therapy, the challenge of achieving consensus treatment targets, and the cumulative cardiovascular risk associated with lifelong LDL-C exposure.^12 6^

Given all trials included patients who had failed to reach LDL-C target of (>3.4 mmol/L [130 mg/dL] at baseline and required patients to be on stable statins/ezetimibe-based therapy for at least 30 days, it is unclear why as many as 17% of participants on placebo achieved an LDL-C reduction to below 130mg/dL. This might partly be explained by natural variations in LDL-C, and inclusion in a trial setting with closer patient monitoring may have improved patients’ adherence to background LLT, although there was insufficient evidence to assess this.

### Implications for practice

Globally, many individuals with FH remain undiagnosed and undertreated, with substantial disparities in access to care between regions and healthcare systems. ^1 2 29 30^ The majority of paediatric patients with HeFH can be managed effectively with statins, with or without ezetimibe, and will not require additional therapies. PCSK9 inhibitors are therefore most relevant for those patients who continue to have substantially elevated LDL-C concentrations despite optimised statins/ezetimibe therapy.

All currently available PCSK9 inhibitors require subcutaneous administration. Among these agents, inclisiran may offer practical advantages because its twice-yearly maintenance dosing schedule.

Given the latest EAS Paediatric FH guideline target of lowering LDL-C to 3.0mmol/l,^6^ the question arises as to what proportion of FH patients under the age of 18 years may fail to achieve this target using only statin/ezetimibe.

Based on published data, rosuvastatin lowers LDL-C by around 40% and the addition of ezetimibe lowers by a further around 15%, resulting in a combined mean LDL-C lowering effect of approximately 55%.^31 21, 32^ It can be estimated that many of those with a pretreatment LDL-C of 6.7 mmol/L and above will not achieve the 3.0mmol/l treatment goal on rosuvastatin treatment alone. For this group, the additional 15% LDL-C lowering likely with ezetimibe will also not bring them to target, and therefore for this group the guideline (Figure 3) recommends instead the use of a PCSK9 inhibitor which can achieve a further 35% reduction. In the published Dutch cohort of children with FH 16.7% had baseline LDL-C of above 6.7mmol/l,^33^ in the UK Paediatric FH Register ^34^ this was 18.4% and in the FHSC paediatric cohort this proportion was around 15%.^35^ Overall, this suggests that around 1 in 6 individuals under 18 years of age who are already being managed for HeFH with standard of care therapy may require a PCSK9i agent in order to achieve the recommended LDL-C targets.

### Future research

Open-label extension studies show encouraging evidence supporting the sustained efficacy and safety of evolocumab and inclisiran for up to two years.^26 14^ However, longer-term follow-up data remain limited and ongoing surveillance is needed to evaluate long-term safety and effectiveness in routine clinical practice.

Although approved by NICE for use in adults with FH who have LDL-C concentrations above target, ^36 37 38^ it is unknown whether or in which specific paediatric FH populations PCSK9 inhibitors may be cost-effective.^39^ These therapies are currently substantially more expensive than established LLTs, and existing economic evaluations have focused almost exclusively on adult populations. ^40 41 42^

Future systematic reviews should incorporate emerging lipid-lowering therapies. Lerodalcibep, a small binding protein that inhibits PCSK9, has been evaluated in adults and in children aged ≥10 years with HoFH and there are plans to investigate it in a phase III trial of children with HeFH aged ≥6 years.^43^ The oral PCSK9 inhibitor enlicitide is currently undergoing phase II evaluation in paediatric populations.^44^

## Conclusions

PCSK9i add-on therapy leads to clinically significant reductions in LDL-C in paediatric patients with HeFH. PCSK9i were generally well tolerated in the short term, with no evidence that PCSK9i have a detrimental effect on growth and maturation. Similar rates of adverse events were observed overall compared with placebo, although analyses were not sufficiently powered to estimate the risk of specific adverse effects. The findings of this systematic review broadly support the 2026 EAS Consensus Statement for the use of PCSK9i in a subset of HeFH children with high LDL-C concentrations despite statin monotherapy. The limited number of current trials with follow-up duration mean that future high-quality studies are needed to further evaluate the long-term effectiveness, tolerability and safety, and cost-effectiveness of PCSK9 inhibitors in this population.

### Financial support and disclaimer

This project was funded by the UK National Institute for Health and Care Research [NIHR134993]. The views expressed are those of the authors and not necessarily those of the NIHR or the Department of Health and Social Care. SEH was supported by a grant from the British Heart Foundation (BHF grant PG 08/008).

## Data Availability

All data produced in the present work are contained in the manuscript.

## Declaration of generative AI use

During the preparation of this manuscript, the first author used Gemini 3.5 Flash to check for concision and accuracy of specific sentences in the background and discussion. After using this tool, the author reviewed and edited the content carefully and takes full responsibility for the content of the published article.

## Acknowledgments

We thank Dr Ralph Akyea, Dr Manpreet Bains, Dr Rebecca Bell-Williams, Prof Joe Kai and Dr Yana Vinogradova, all from the Centre for Academic Primary Care, University of Nottingham, UK, for contributions during the inception of this review, its conduct and/or the dissemination of its findings.

