## Supplementary file for "Efficacy and safety of PCSK9 inhibitors for children and adolescents with heterozygous familial hypercholesterolaemia: Systematic review and meta-analysis of randomised controlled trials"

### Contents

Risk of bias assessment (RoB 2) ... 2
Supplementary tables ... 2
Supplementary figures ... 5
Literature search and search strategies ... 7

#### Supplementary tables

Table 1 HeFH diagnosis method and trial selection criteria ... 2
Table 2 Mean absolute change in LDL cholesterol from baseline (mg/dl or percentage) ... 3
Table 3 Mean absolute change in LDL cholesterol from baseline (mmol/l or percentage) ... 3
Table 4 LDL-C target outcomes (other than 130mg/dl) ... 3
Table 5 LS mean % change in HDL-C from baseline ... 4
Table 6 LS mean % change in triglycerides from baseline ... 4
Table 7 Laboratory safety endpoints ... 4

#### Supplementary figures

Figure 1 Study selection process (PRISMA flow diagram) ... 5
Figure 2 LS mean change in absolute LDL-C (mg/dl) ... 6
Figure 3 LS mean percentage change from baseline in total cholesterol ... 6
Figure 4 Mean difference in change from baseline in weight (kg) ... 6
Figure 5 Mean difference in change from baseline in height (cm) ... 7
Figure 6 Serious adverse event ... 7
Figure 7 Injection site reactions ... 7

#### Risk of bias assessment (RoB 2)

| Study ID | D1 | D2 | D3* | D4 | D5 | Overall |
|---|---|---|---|---|---|---|
| Santos 2020 (HAUSER-RCT) | + | + | ! | + | + | + |
| Santos 2024 | + | + | + | + | + | + |
| Wiegman 2026 (ORION-16) | + | + | + | + | + | + |

+ Low risk
! Some concerns
- High risk

D1: randomisation process; D2: deviation from intended interventions; D3: missing outcome data; D4: measurement of the outcome; D5: selection of the reported result.

All risk of bias assessments were conducted for the least-squares mean differences in % reduction in LDL-C outcome. In Santos (2020), outcome data at week 24 were missing for 17% of participants in the placebo arm and 8% in the evolocumab arm. Reasons for missing outcome data by group and results of a sensitivity analysis with multiple imputation were not reported. Although a mixed model for repeated measures was used, the plausibility of the missing-at-random assumption was uncertain. Therefore, bias due to missing outcome data was considered to raise some concerns. The expected magnitude of any bias would likely be small (and may favour placebo). Therefore, overall risk of bias for Santos (2020) was considered low.

#### Supplementary tables

*Table 1 HeFH diagnosis method and trial selection criteria*

| | Diagnosis | Selection criteria |
|---|---|---|
| HAUSER-RCT | Genetic testing or Simon Broome (definite & possible), Dutch Lipid Clinic Network (definite and probable) or Make Early Diagnosis to Prevent Early Deaths (MEDPED) | **Inclusion**<br>10 - 17 years; HeFH, ≥LDL-C 130 mg/dL (3.4 mmol/L) & TG ≤400 mg/dL (4.5 mmol/L). Stable optimised LLT for ≥4 wks prior to screening as per local guidelines, not requiring up titration in the opinion of the managing physician. SoC required to remain stable during the study.<br><br>**Exclusion**<br>HoFH, diabetes, abnormal thyroid levels, abnormal free thyroxine, AST/ALT>2x normal, creatine kinase>3x normal, pregnancy, others. |
| Santos (2024) | Genetic testing (90%) or Simon Broome | **Inclusion**<br>8 - 17 years, HeFH, ≥LDL-C 130 mg/dL (3.4 mmol/L). Optimal stable LLT for ≥4 wks prior to screening defined according to regional practice or local guidelines, or as maximally tolerated statin dose due to adverse effects of higher doses. SoC required to remain stable during the study.<br><br>**Exclusion**<br>25 kg in body weight, aged 8 to 9 years and not at Tanner Stage 1, or aged 10 to 17 years and not at Tanner Stage 2 or higher, secondary hyperlipidemia; HoFH; prior lipid apheresis within 2 months of screening or planned apheresis during the study; uncontrolled type 1 or 2 diabetes (per local guidelines), thyroid disease or hypertension; |

| | | |
|---|---|---|
| | | severe kidney impairment; abnormal liver function; or creatine phosphokinase greater than 3 times the upper limit of normal. |
| ORION-16 | Genetic testing and/or clinical | **Inclusion**<br>12 - <18 years, LDL-C>3.4 mmol/L (130 mg/dL) on stable maximally tolerated/optimal statin +/- other LLT for ≥4 wks prior to screening. Optimal SoC defined according to international and local practice, treatment guidelines and regulatory bodies. SoC required to remain stable during the study (up to Year 1).<br><br>**Exclusion**<br>HoFH, active liver disease, secondary hypercholesterolaemia (eg, hypo thyroidism or nephrotic syndrome), previous treatment with monoclonal antibodies targeting PCSK9 within 90 days of screening, or planned use of other investigational medicinal products or devices. |

*Table 2 Mean absolute change in LDL cholesterol from baseline (mg/dl or percentage)*

| Treatment | Follow-up | PCSK9i* | Placebo* | LS mean difference in absolute change from baseline between PCSK9i and placebo* |
|---|---|---|---|---|
| Evolocumab | 24 weeks | -77.5 (−86.1 to -68.9) | -9.0 (−21.1 to 3.2) | -68.6 (−83.1 to −54.0) |
| Alirocumab-Q4W | 24 weeks | -66.6 (SD 7.7) | -15.3 (SD 7) | -33.8% (-46.4 to -21.2) |
| Alirocumab-Q2W | 24 weeks | -58.9 (SD 5.3) | 14.5 (SD 8.2) | -43.3% (97.5% CI -56.0 to -30.7) |
| Inclisiran[#] | 47 weeks | -50.27 (-58.01 to -42.54) | 0 (-11.60 to 7.73) | -50.27 (-61.87 to -38.67) |

* Intervals in brackets are 95% CI.
[#] Values converted from mmol/L

*Table 3 Mean absolute change in LDL cholesterol from baseline (mmol/l or percentage)*

| Treatment | Follow-up | PCSK9i* | Placebo* | LS mean difference in absolute change from baseline between PCSK9i and placebo* |
|---|---|---|---|---|
| Evolocumab[#] | 24 weeks | −2.00 (−2.23 to −1.78) | −0.23 (−0.55 to 0.08) | −1.77 (−2.15 to −1.40) |
| Alirocumab-Q4W[#] | 24 weeks | −1.72 (SD 0.20) | −0.40 (SD 0.18) | -33.8% (-46.4 to -21.2) |
| Alirocumab-Q2W[#] | 24 weeks | −1.52 (SD 0.14) | 0.37 (SD 0.21) | -43.3% (97.5% CI -56.0 to -30.7) |
| Inclisiran | 47 weeks | -1.3 (-1.5 to -1.1) | 0 (-0.3 to 0.2) | -1.3 (-1.6 to -1.0) |

* Intervals in brackets are 95% CI, unless otherwise specified.
[#] Values converted from mg/dl

*Table 4 LDL-C target outcomes (other than 130mg/dl)*

| Treatment | Follow-up | PCSK9i* | Placebo* | PCSK9i vs. placebo |
|---|---|---|---|---|
| ***Reaching <110 mg/dl (2.84 mmol/L)*** | | | | |
| Alirocumab Q4W | 24 weeks | NR | NR | OR 43.1 (97.5% 3.7 to 498.6) |
| Alirocumab Q2W | 24 weeks | NR | NR | OR 52.7 (97.5% 3.5 to 804.3) |
| Inclisiran | 47 weeks | 27% (25/93) | 0 (0/48) | NR |

| | | | | |
|---|---|---|---|---|
| ***Reaching <100mg/dl (2.59 mmo/L)*** | | | | |
| Evolocumab | 24 weeks | 58% (60/104) | 2% (1/53) | NR |
| ***Reduction of >50% from baseline*** | | | | |
| Evolocumab | 24 weeks | 41% (43/104) | 2% (1/53) | NR |

*Rates are calculated based on the ITT principle

*Table 5 LS mean % change in HDL-C from baseline*

| **Treatment** | **Follow-up** | **PCSK9i** | **Placebo** | **PCSK9i vs. placebo** |
|---|---|---|---|---|
| Alirocumab Q4W | 24 weeks | NR | NR | 4.4% (97.5% −3.6 to 12.5) |
| Alirocumab Q2W | 24 weeks | NR | NR | 6.4% (97.5% 0.5 to 12.3) |
| Inclisiran | 47 weeks | 7.40% (SD 19.1) | -1.90% (16.1) | NR |

*Table 6 LS mean % change in triglycerides from baseline*

| **Treatment** | **Follow-up** | **PCSK9i** | **Placebo** | **PCSK9i vs. placebo** |
|---|---|---|---|---|
| Alirocumab Q4W | 24 weeks | NR | NR | -19.0% (97.5% -41.5 to 3.5) |
| Alirocumab Q2W | 24 weeks | NR | NR | 4.3% (97.5% -19.1 to 27.6) |
| Inclisiran | 47 weeks | 1.8% (SD 38.4) | 9.7% (SD 43.0) | NR |

*Table 7 Laboratory safety endpoints*

| | **Treatment** | **Follow-up** | **PCSK9i** | **Placebo** |
|---|---|---|---|---|
| **Creatine kinase >5 × ULN** | Evolocumab | 24 weeks | 0 | 0 |
| **Aminotransferase >3 x ULN** | Evolocumab | 24 weeks | 0 | 0 |
| **Creatine kinase >5 × ULN** | Inclisiran | 47 weeks | 3 (3%) | 3 (6%) |
| **Creatinine >2 mg/dL** | Inclisiran | 47 weeks | 0 | 0 |
| **ALT >3 × ULN** | Inclisiran | 47 weeks | 0 | 0 |
| **Total bilirubin >2 × ULN** | Inclisiran | 47 weeks | 3 (3%) | 1 (2%) |

#### Supplementary figures

*Figure 1 Study selection process (PRISMA flow diagram)*

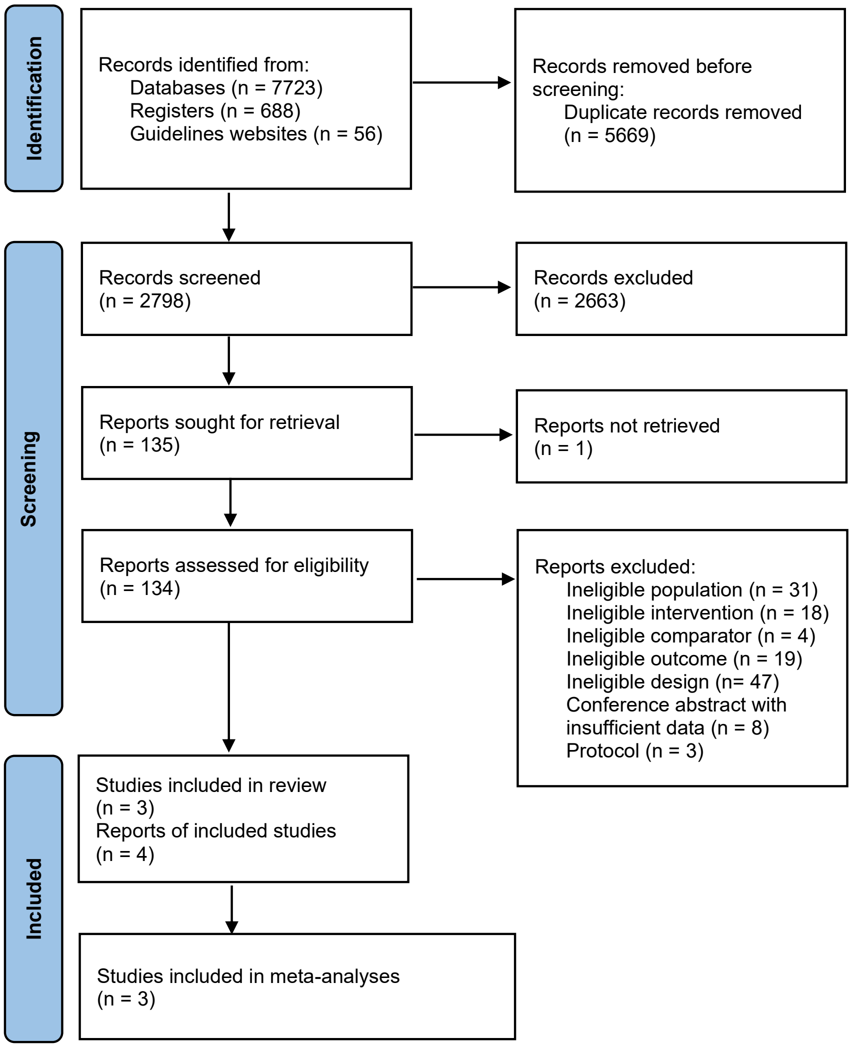

*Figure 2 LS mean change in absolute LDL-C (mg/dl)*

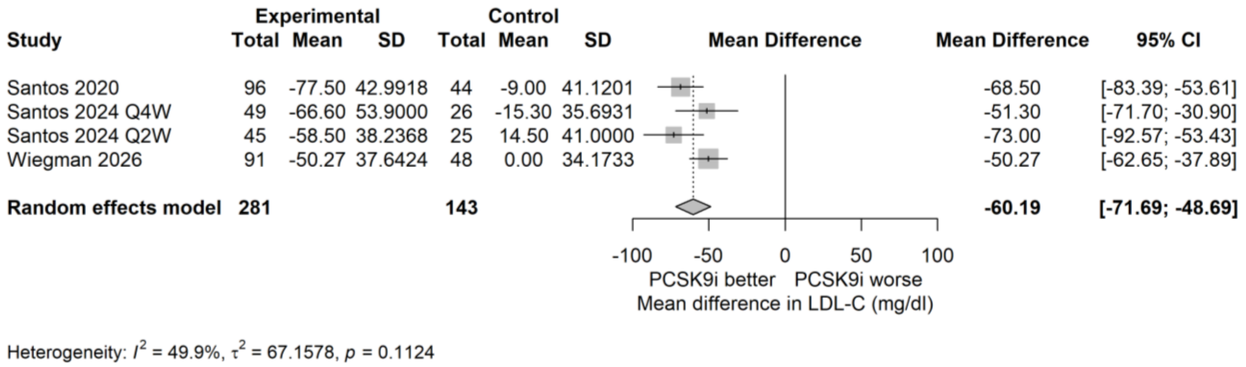


*Figure 3 LS mean percentage change from baseline in total cholesterol*

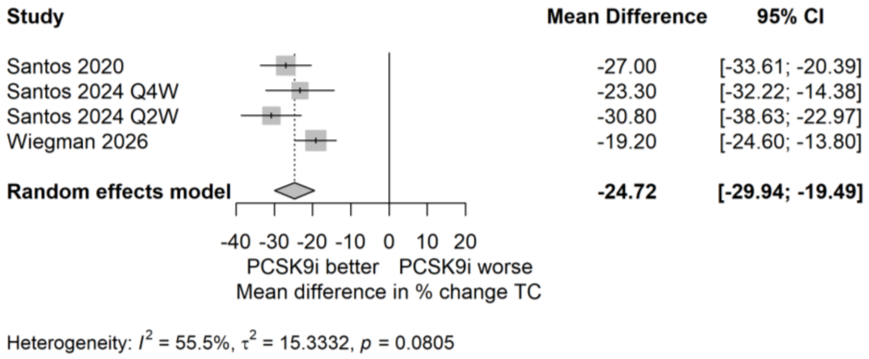


*Figure 4 Mean difference in change from baseline in weight (kg)*

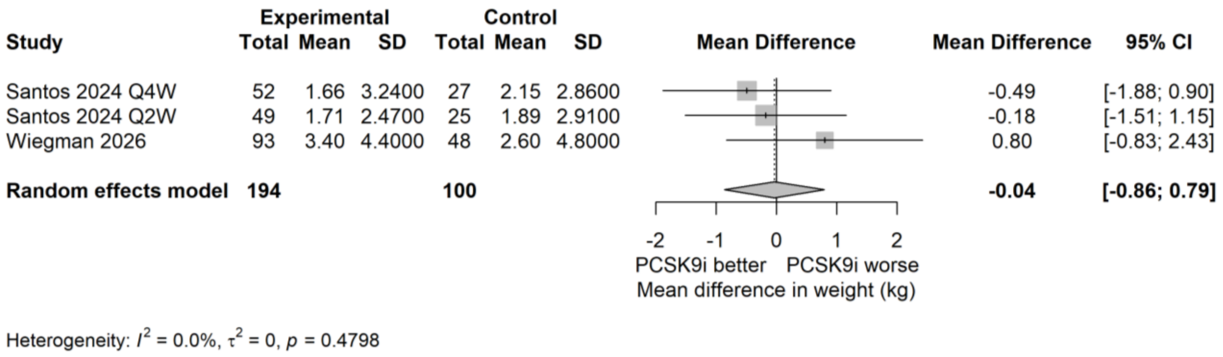

*Figure 5 Mean difference in change from baseline in height (cm)*

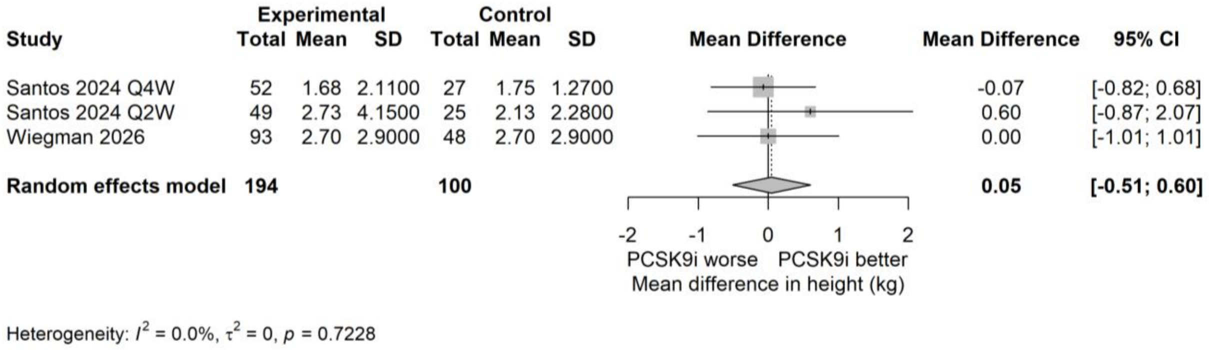


*Figure 6 Serious adverse event*

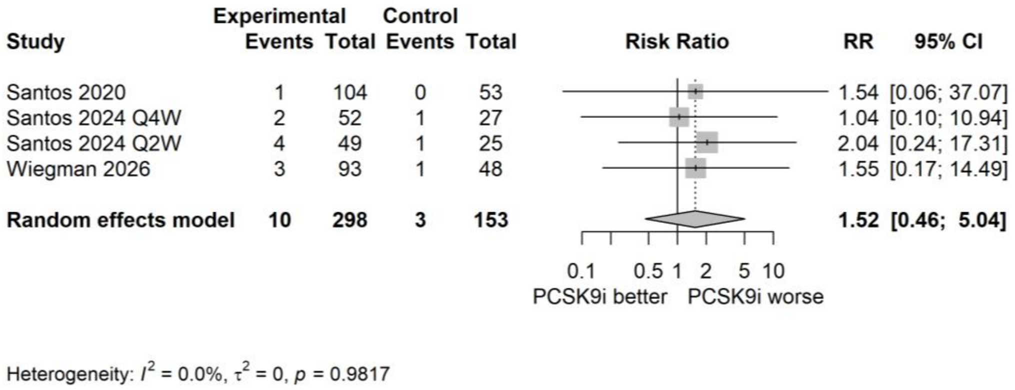


*Figure 7 Injection site reactions*

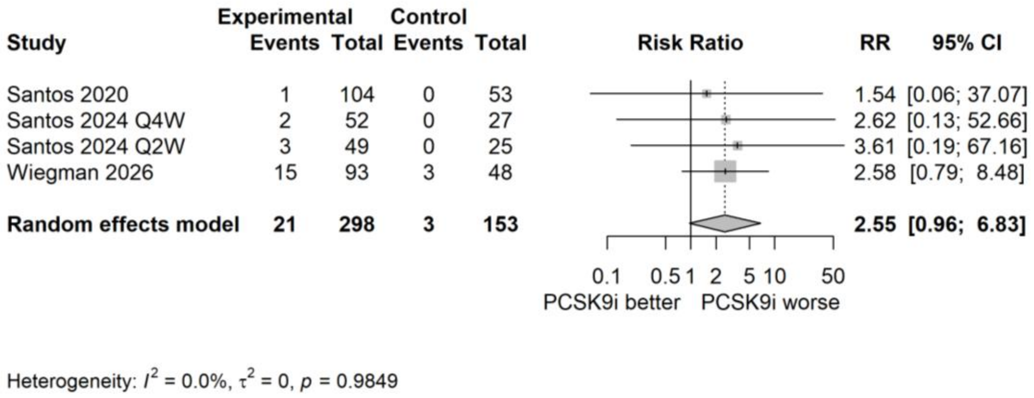


#### Literature search and search strategies

The aim of the search was to systematically identify published and unpublished studies of lipid-lowering therapies in children aged 0-18 years with heterozygous familial hypercholesterolaemia. The Ovid MEDLINE search strategy for a previous review by the same author team was updated, with the intervention terms expanded to include PCSK9-targeted therapies.[1]

The strategy was structured using the following three concepts:
1. children (0-18 years)
AND
2. heterozygous familial hypercholestrolaemia
AND
3. lipid-lowering therapies (statins, ezetimibe, and PCSK9-targeted therapies)

Search terms for each concept were identified through examination of terms used in key papers, online drug information resources, use of database thesauri and through discussion with the review team. Relevant subject headings and textword searches of the title and abstracts of records were utilised in the strategy. Retrieval was not restricted by language, date or study design.

The strategy was peer reviewed by a second information specialist using aspects of the PRESS checklist.[2] The final MEDLINE strategy was adapted for use in all resources searched.

The following databases were searched in February 2026:

- MEDLINE ALL (Ovid)
- Cochrane Controlled Register of Trials (Wiley)
- Cochrane Database of Systematic Reviews (Wiley)
- Embase (Ovid)
- International Health Technology Assessment (INAHTA) database
- Science Citation Index (Web of Science)

Further ongoing and unpublished studies were identified through searches of:

- ClinicalTrials.gov
- Conference Proceedings Citation Index: Science (Web of Science)
- EU Clinical Trials Register
- PROSPERO
- WHO International Clinical Trials Registry Platform portal

A search for relevant guidelines was carried out via the following websites: National Institute for Health and Clinical Excellence (NICE), Guidelines International Network (GIN) and the Trip database. Supplementary searching of the reference lists of relevant reviews and included studies was undertaken to identify any additional relevant studies.

All search results were imported into the original EndNote library for the previous review.[1] Any duplicates with records identified for the previous review were identified and removed.

Full search strategies for all resources can be found below.

***Database search strategies***

**MEDLINE ALL**
(includes: Epub Ahead of Print, In-Process & Other Non-Indexed Citations, Ovid MEDLINE Daily and Ovid MEDLINE)
via Ovid http://ovidsp.ovid.com/
Date range: 1946 to February 24, 2026
Date searched: 25$^{th}$ February 2026

Records retrieved: 1893

1 Hyperlipoproteinemia Type II/ (8237)
2 hyperlipidemias/ (29551)
3 hypercholesterolemia/ (28036)
4 (hyperlipoprotein?emi$ or (hyper adj3 lipoprotein?emi$) or hyperbetalipoprotein?emi$).ti,ab. (4798)
5 (hypercholesterol?emi$ or hyper cholesterol?emi$).ti,ab. (42235)
6 (hyperlipid?emi$ or hyper lipid?emi$).ti,ab. (41813)
7 heFH.ti,ab. (407)
8 heterozygous FH.ti,ab. (612)
9 1 or 2 or 3 or 4 or 5 or 6 or 7 or 8 (111220)
10 exp Child/ (2315057)
11 Adolescent/ (2387663)
12 exp Infant/ (1334972)
13 (child or children or childhood$ or infant$ or infancy or pediatric$ or paediatric$ or preschool$ or pre school$ or schoolchild$ or school age$ or schoolage$ or schoolboy$ or schoolgirl$).ti,ab. (2384320)
14 (girl or girls or boy or boys or kid or kids).ti,ab. (304317)
15 (adolesc$ or young people or young person$ or teen$ or youth$ or preteen$ or pubert$ or prepubert$ or pubescen$ or prepubescen$ or juvenil$).ti,ab. (681687)
16 (neonat$ or neo nat$ or newborn$ or new born$ or newly born$ or baby or babies).ti,ab. (552252)
17 or/10-16 (5284851)
18 9 and 17 (12447)
19 exp Hydroxymethylglutaryl-CoA Reductase Inhibitors/ (50207)
20 Atorvastatin/ (7986)
21 Fluvastatin/ (1495)
22 Lovastatin/ (4848)
23 Pravastatin/ (3610)
24 Rosuvastatin Calcium/ (3252)
25 exp Simvastatin/ (8798)
26 (HMG-CoA or hydroxymethylglutaryl CoA reductase inhibitor$ or hydroxymethylglutaryl coenzyme a inhibitor$).af. (42997)
27 (atorvastatin$ or lipitor$ or lypqozet$ or caduet$).af. (12758)
28 (fluvastatin$ or lescol$ or nandovar$ or dorisin$ or fluindostatin$).af. (2381)
29 (lovastatin$ or mevacor$ or altoprev$ or mevinolin$).af. (6684)
30 (pitavastatin$ or livalo$ or zypitamag$).af. (1369)
31 (pravastatin$ or pravachol$).af. (5363)
32 (rosuvastatin$ or crestor$ or ezallor$).af. (5395)
33 (simvastatin$ or zocor$ or flolipid$ or vytorin$ or inegy$).af. (13130)
34 statin$.af. (60482)
35 or/19-34 (90466)
36 18 and 35 (1282)
37 exp Ezetimibe/ (2864)
38 (ezetimibe$ or ezetrol$ or zetia$).af. (5759)
39 37 or 38 (5759)
40 18 and 39 (286)
41 ((lipid-modif$ or lipid-lower$) adj3 (therap$ or treatment$ or intervention$ or medication$ or drug$ or agent$ or inject$)).ti,ab. (16012)
42 18 and 41 (525)

43 36 or 40 (1331)
44 42 or 43 (1602)
45 PCSK9 Inhibitors/ (1609)
46 ((PCSK9$ adj3 (inhibitor$ or block$ or antagonist$ or therap$ or target$)) or PCSKi$).af. (3899)
47 ((proprotein convertase subtilisin or kexin type 9) adj3 (inhibitor$ or block$ or antagonist$)).af. (1558)
48 antibodies, monoclonal/ or antibodies, monoclonal, humanized/ (248231)
49 (monoclonal antibod$ or mAb$).af. (277694)
50 (Evolocumab$ or repatha$ or AMG-145 or AMG145 or sal-003 or sal003).af. (1461)
51 (Alirocumab$ or praluent$ or SAR-236553 or SAR236553 or REGN727 or REGN-727).af. (1212)
52 (enlicitide$ or MK-0616 or MK0616).af. (26)
53 (Inclisiran$ or leqvio$ or ALN-PCS$ or ALNPCS$ or aln 60212 or aln60212 or kjx 839 or kjx839).af. (643)
54 RNA, Small Interfering/ (76653)
55 ((small or short) adj2 (interfering RNA or interfering ribonucleic acid)).af. (32816)
56 siRNA$.af. (87929)
57 bile acid sequest$.af. (924)
58 Colesevelam Hydrochloride/ (233)
59 (colesevelam$ or cholestagel$ or lodalis$ or welchol or gt31 104$ or gt 31 104$ or iw 3718 or iw3718).af. (381)
60 (colestilan$ or bindren$ or cholebine$ or colestimide$ or mci 196 or mci196).af. (107)
61 Colestipol/ (399)
62 (colestipol$ or colestid$ or cholestabyl$ or lestid$).af. (634)
63 Cholestyramine Resin/ (2696)
64 (cholestyramin$ or colestyramin$ or cuemid$ or quantalan$ or questran$ or olestyr$ or prevalite$ or mk 135 or mk135).af. (3805)
65 DEAE-Dextran/ (471)
66 (diethylaminoethyldextran$ or diethylaminoethyl-dextran$ or DEAE-dextran$ or DEAE-sephadex$ or pulsar$ or dexide$ or nolipid$).af. (4508)
67 (bempedoic acid$ or bempedoate$ or nexleto$ or nilemdo$ or nexlizet$ or nustendi$ or esp 55016 or esp55016 or etc 1002 or etc1002).af. (641)
68 (lerodalcibep$ or lerochol$ or "lib 003" or lib003).af. (14)
69 or/45-68 (547110)
70 18 and 69 (566)
71 44 or 70 (1905)
72 exp animals/ not humans/ (5427820)
73 71 not 72 (1893)

**Key:**
/ = subject heading (MeSH heading)
exp = exploded subject heading (MeSH heading)
$ = truncation
? = optional wild card character, stands for zero or one character within a word
ti,ab = terms in title or abstract fields
af = terms in any fields
adj3 = terms within three words of each other (any order)

**Cochrane Controlled Register of Trials (CENTRAL)**
via Wiley http://onlinelibrary.wiley.com/

Issue: Issue 1 of 12, January 2026
Date searched: 25th February 2026
Records retrieved: 465

#1 MeSH descriptor: [Hyperlipoproteinemia Type II] this term only 672
#2 MeSH descriptor: [Hypercholesterolemia] this term only 4260
#3 MeSH descriptor: [Hyperlipidemias] this term only 2479
#4 (hyperlipoprotein*mi* or (hyper near/3 lipoprotein*emi*) or hyperbetalipoprotein*mi*):ti,ab,kw 1650
#5 (hypercholesterol*mi* or hyper cholesterol*mi*):ti,ab,kw 9310
#6 (hyperlipid*mi* or hyper lipid*mi*):ti,ab,kw 7897
#7 heFH:ti,ab,kw 204
#8 heterozygous FH:ti,ab,kw 300
#9 #1 or #2 or #3 or #4 or #5 or #6 or #7 or #8 16630
#10 MeSH descriptor: [Child] explode all trees 84328
#11 MeSH descriptor: [Adolescent] this term only 139881
#12 MeSH descriptor: [Infant] explode all trees 46714
#13 (child or children or childhood* or infant* or infancy or pediatric* or paediatric* or preschool* or (pre next school*) or schoolchild* or (school next age*) or schoolage* or schoolboy* or schoolgirl*):ti,ab,kw 260626
#14 (girl or girls or boy or boys or kid or kids):ti,ab,kw 15977
#15 (adolesc* or (young next people) or (young next person*) or teen* or youth* or preteen* or pubert* or prepubert* or pubescen* or prepubescen* or juvenil*):ti,ab,kw 191505
#16 (neonat* or neo-nat* or newborn* or new-born* or (newly next born*) or baby or babies):ti,ab,kw 58054
#17 #10 or #11 or #12 or #13 or #14 or #15 or #16 398435
#18 #9 and #17 1236
#19 MeSH descriptor: [Hydroxymethylglutaryl-CoA Reductase Inhibitors] explode all trees 5077
#20 MeSH descriptor: [Atorvastatin] this term only 2970
#21 MeSH descriptor: [Fluvastatin] this term only 377
#22 MeSH descriptor: [Lovastatin] this term only 687
#23 MeSH descriptor: [Pravastatin] this term only 1225
#24 MeSH descriptor: [Rosuvastatin Calcium] this term only 1453
#25 MeSH descriptor: [Simvastatin] explode all trees 2173
#26 ((atorvastatin* or lipitor* or lypqozet* or caduet*)) 6514
#27 ((fluvastatin* or lescol* or nandovar* or dorisin* or fluindostatin*)) 846
#28 ((lovastatin* or mevacor* or altoprev* or mevinolin*)) 1122
#29 ((pitavastatin* or livalo* or zypitamag*)) 657
#30 ((pravastatin* or pravachol*)) 2149
#31 ((rosuvastatin* or crestor* or ezallor*)) 3217
#32 ((simvastatin* or zocor* or flolipid* or vytorin* or inegy*)) 4392
#33 ((HMG-CoA or ("hydroxymethylglutaryl CoA reductase" next inhibitor*) or ("hydroxymethylglutaryl coenzyme a" next inhibitor*))) 46
#34 (statin*) 13678
#35 #19 or #20 or #21 or #22 or #23 or #24 or #25 or #26 or #27 or #28 or #29 or #30 or #31 or #32 or #33 or #34 22596
#36 #18 and #35 371
#37 MeSH descriptor: [Ezetimibe] explode all trees 1035
#38 ((ezetimibe* or ezetrol* or zetia*)) 2194
#39 #37 or #38 2194

#40 #18 and #39 90
#41 ((lipid-modif* or lipid-lower*) near/2 (therap* or treatment* or intervention* or medication* or drug* or agent* or inject*)) 3765
#42 #18 and #41 114
#43 MeSH descriptor: [PCSK9 Inhibitors] this term only 218
#44 ((PCSK9* near/3 (inhibitor* or block* or antagonist* or therap* or target*)) or PCSKi*):ti,ab,kw 738
#45 (("proprotein convertase subtilisin" or "kexin type 9") near/3 (inhibitor* or block* or antagonist*)):ti,ab,kw 269
#46 MeSH descriptor: [Antibodies, Monoclonal] this term only 7946
#47 MeSH descriptor: [Antibodies, Monoclonal, Humanized] this term only 8401
#48 (monoclonal next antibod* or mAb*):ti,ab,kw 16529
#49 (Evolocumab* or repatha* or AMG-145 or AMG145 or sal-003 or sal003):ti,ab,kw 619
#50 (Alirocumab* or praluent* or SAR-236553 or SAR236553 or REGN727 or REGN-727):ti,ab,kw 527
#51 (enlicitide* or MK-0616 or MK0616):ti,ab,kw 33
#52 (Inclisiran* or leqvio* or ALN-PCS* or ALNPCS* or aln-60212 or aln60212 or kjx-839 or kjx839):ti,ab,kw 167
#53 MeSH descriptor: [RNA, Small Interfering] this term only 166
#54 ((small or short) near/2 (interfering next RNA or "interfering ribonucleic acid")):ti,ab,kw 227
#55 siRNA*:ti,ab,kw 339
#56 bile next acid next sequest*:ti,ab,kw 184
#57 MeSH descriptor: [Colesevelam Hydrochloride] this term only 129
#58 (colesevelam* or cholestagel* or lodalis* or welchol or gt31 104* or gt 31 104* or iw 3718 or iw3718):ti,ab,kw 224
#59 (colestilan$ or bindren$ or cholebine$ or colestimide$ or mci-196 or mci196):ti,ab,kw 56
#60 MeSH descriptor: [Colestipol] this term only 109
#61 (colestipol* or colestid* or cholestabyl* or lestid*):ti,ab,kw 164
#62 MeSH descriptor: [Cholestyramine Resin] this term only 317
#63 (cholestyramin* or colestyramin* or cuemid* or quantalan* or questran* or olestyr* or prevalite* or mk-135 or mk135):ti,ab,kw 531
#64 MeSH descriptor: [DEAE-Dextran] this term only 8
#65 (diethylaminoethyldextran* or diethylaminoethyl-dextran* or DEAE-dextran* or DEAE-sephadex* or pulsar* or dexide* or nolipid*):ti,ab,kw 126
#66 (bempedoic next acid* or bempedoate* or nexleto* or nilemdo* or nexlizet* or nustendi* or esp-55016 or esp55016 or etc-1002 or etc1002):ti,ab,kw 213
#67 (lerodalcibep* or lerochol* or lib-003 or lib003):ti,ab,kw 28
#68 #43 or #44 or #45 or #46 or #47 or #48 or #49 or #50 or #51 or #52 or #53 or #54 or #55 or #56 or #57 or #58 or #59 or #60 or #61 or #62 or #63 or #64 or #65 or #66 or #67 29063
#69 #18 and #68 147
#70 #36 or #40 or #42 or #69 in Cochrane Reviews, Cochrane Protocols 7
#71 #36 or #40 or #42 or #69 in Trials 465

**Key:**
MeSH descriptor = subject heading (MeSH heading)
* = truncation
ti,ab,kw = terms in title, abstract or keyword fields
near/3 = terms within three words of each other (any order)
next = terms are next to each other

**Cochrane Database of Systematic Reviews (CDSR)**
via Wiley http://onlinelibrary.wiley.com/
Issue: Issue 2 of 12, February 2026
Date searched: 25th February 2026
Records retrieved: 7

See above under CENTRAL for search strategy.

**Embase**
via Ovid http://ovidsp.ovid.com/
Date range: 1974 to 2026 February 23
Date searched: 25th February 2026
Records retrieved: 2960

1 familial hypercholesterolemia/ (16302)
2 hypercholesterolemia/ (83870)
3 hyperlipidemia/ (116438)
4 (hyperlipoprotein?emi$ or (hyper adj3 lipoprotein?emi$) or hyperbetalipoprotein?emi$).ti,ab. (5651)
5 (hypercholesterol?emi$ or hyper cholesterol?emi$).ti,ab. (63180)
6 (hyperlipid?emi$ or hyper lipid?emi$).ti,ab. (72345)
7 heFH.ti,ab. (905)
8 heterozygous FH.ti,ab. (986)
9 or/1-8 (234978)
10 exp child/ (3556609)
11 exp adolescence/ (110655)
12 exp infant/ (1277472)
13 juvenile/ (63466)
14 (child or children or childhood$ or infant$ or infancy or pediatric$ or paediatric$ or preschool$ or pre school$ or schoolchild$ or school age$ or schoolage$ or schoolboy$ or schoolgirl$).ti,ab. (3122187)
15 (girl or girls or boy or boys or kid or kids).ti,ab. (413290)
16 (adolesc$ or young people or young person$ or teen$ or youth$ or preteen$ or pubert$ or prepubert$ or pubescen$ or prepubescen$ or juvenil$).ti,ab. (899018)
17 (neonat$ or neo nat$ or newborn$ or new born$ or newly born$ or baby or babies).ti,ab. (724638)
18 or/10-17 (5079634)
19 9 and 18 (16115)
20 exp hydroxymethylglutaryl coenzyme A reductase inhibitor/ (223674)
21 atorvastatin/ (54120)
22 atorvastatin plus ezetimibe/ (222)
23 amlodipine plus atorvastatin/ (397)
24 fluindostatin/ (11257)
25 mevinolin/ (18244)
26 pravastatin/ (22934)
27 rosuvastatin/ (23776)
28 simvastatin/ (46572)
29 ezetimibe plus simvastatin/ (1654)
30 (HMG-CoA or hydroxymethylglutaryl CoA reductase inhibitor$ or hydroxymethylglutaryl coenzyme a inhibitor$).af. (14917)

31 (atorvastatin$ or lipitor$ or lypqozet$ or caduet$).af. (55477)
32 (fluvastatin$ or lescol$ or nandovar$ or dorisin$ or fluindostatin$).af. (11471)
33 (lovastatin$ or mevacor$ or altoprev$ or mevinolin$).af. (18992)
34 (pitavastatin$ or livalo$ or zypitamag$).af. (5162)
35 (pravastatin$ or pravachol$).af. (23424)
36 (rosuvastatin$ or crestor$ or ezallor$).af. (24348)
37 (simvastatin$ or zocor$ or flolipid$ or vytorin$ or inegy$).af. (48458)
38 statin$.af. (109299)
39 or/20-38 (259422)
40 19 and 39 (2389)
41 ezetimibe/ (17029)
42 (ezetimibe$ or ezetrol$ or zetia$).af. (19416)
43 41 or 42 (19416)
44 19 and 43 (751)
45 ((lipid-modif$ or lipid-lower$) adj2 (therap$ or treatment$ or intervention$ or medication$ or drug$ or agent$ or inject$)).ti,ab. (25652)
46 19 and 45 (635)
47 40 or 44 (2455)
48 46 or 47 (2718)
49 PCSK9 inhibitor/ (2733)
50 ((PCSK9$ adj3 (inhibitor$ or block$ or antagonist$ or therap$ or target$)) or PCSKi$).af. (6872)
51 ((proprotein convertase subtilisin or kexin type 9) adj3 (inhibitor$ or block$ or antagonist$)).af. (2302)
52 monoclonal antibody/ (252424)
53 (monoclonal antibod$ or mAb$).af. (477499)
54 evolocumab/ (4499)
55 (Evolocumab$ or repatha$ or AMG-145 or AMG145 or sal-003 or sal003).af. (4711)
56 alirocumab/ (3518)
57 (Alirocumab$ or praluent$ or SAR-236553 or SAR236553 or REGN727 or REGN-727).af. (3678)
58 enlicitide/ (71)
59 (enlicitide$ or "MK 0616" or MK0616).af. (78)
60 inclisiran/ (1598)
61 (Inclisiran$ or leqvio$ or ALN-PCS$ or ALNPCS$ or aln 60212 or aln60212 or kjx 839 or kjx839).af. (1740)
62 small interfering RNA/ (177203)
63 ((small or short) adj2 (interfering RNA or interfering ribonucleic acid)).af. (182976)
64 siRNA$.af. (132622)
65 exp bile acid sequestrant/ (18607)
66 bile acid sequest$.af. (2785)
67 (colesevelam$ or cholestagel$ or lodalis$ or welchol$ or gt31 104$ or gt 31 104$ or iw 3718 or iw3718).af. (1805)
68 (colestilan$ or bindren$ or cholebine$ or colestimide$ or mci 196 or mci196).af. (320)
69 (colestipol$ or colestid$ or cholestabyl$ or lestid$).af. (3154)
70 (cholestyramin* or colestyramin$ or cuemid$ or quantalan$ or questran$ or olestyr$ or prevalite$ or mk 135 or mk135).af. (12220)
71 (diethylaminoethyldextran$ or diethylaminoethyl-dextran$ or DEAE-dextran$ or DEAE-sephadex$ or pulsar$ or dexide$ or nolipid$).af. (7803)
72 bempedoic acid/ (1485)

73 (bempedoic acid$ or bempedoate$ or nexleto$ or nilemdo$ or nexlizet$ or nustendi$ or esp 55016 or esp55016 or etc 1002 or etc1002).af. (1605)
74 lerodalcibep/ (69)
75 (lerodalcibep$ or lerochol$ or "lib 003" or lib003).af. (75)
76 or/49-75 (709323)
77 19 and 76 (1020)
78 48 or 77 (3048)
79 limit 78 to "remove clinical trial (clinicaltrials.gov) records" (2960)

**Key:**
/ = subject heading (Emtree heading)
exp = exploded subject heading (Emtree heading)
$ = truncation
? = optional wild card character, stands for zero or one character within a word
ti,ab = terms in title or abstract fields
af = terms in all fields
adj3 = terms within three words of each other (any order)

**International Health Technology Assessment (INAHTA) database**
via https://database.inahta.org/
Date searched: 26th February 2026
Records retrieved: 11

1. ((((neonat* OR neo-nat* OR newborn* OR new-born* OR "newly born" OR baby OR babies)[Title] OR (neonat* OR neo-nat* OR newborn* OR new-born* OR "newly born" OR baby OR babies)[abs] OR (neonat* OR neo-nat* OR newborn* OR new-born* OR "newly born" OR baby OR babies)[Keywords]) OR ((adolesc* OR "young people" OR "young person" OR teen* OR youth* OR preteen* OR pubert* OR prepubert* OR pubescen* OR prepubescen* OR juvenil*)[Title] OR (adolesc* OR "young people" OR "young person" OR teen* OR youth* OR preteen* OR pubert* OR prepubert* OR pubescen* OR prepubescen* OR juvenil*)[abs] OR (adolesc* OR "young people" OR "young person" OR teen* OR youth* OR preteen* OR pubert* OR prepubert* OR pubescen* OR prepubescen* OR juvenil*)[Keywords]) OR ((girl OR girls OR boy OR boys OR kid OR kids)[Title] OR (girl OR girls OR boy OR boys OR kid OR kids)[abs] OR (girl OR girls OR boy OR boys OR kid OR kids)[Keywords]) OR ((child OR children OR childhood* OR infant* OR infancy OR pediatric* OR paediatric* OR preschool* OR pre-school* OR schoolchild* OR school-age* OR schoolage* OR schoolboy* OR schoolgirl*)[Title] OR (child OR children OR childhood* OR infant* OR infancy OR pediatric* OR paediatric* OR preschool* OR pre-school* OR schoolchild* OR school-age* OR schoolage* OR schoolboy* OR schoolgirl*)[abs] OR (child OR children OR childhood* OR infant* OR infancy OR pediatric* OR paediatric* OR preschool* OR pre-school* OR schoolchild* OR school-age* OR schoolage* OR schoolboy* OR schoolgirl*)[Keywords]) OR ("Infant"[mhe]) OR ("Adolescent"[mh]) OR ("Child"[mhe])) AND (((heFH OR "heterozygous FH")[Title] OR (heFH OR "heterozygous FH")[abs] OR (heFH OR "heterozygous FH")[abs]) OR ((hypercholesterolemi* OR hypercholesterolaemi* OR hyperlipidemi* OR hyperlipidaemi* )[Title] OR (hypercholesterolemi* OR hypercholesterolaemi* OR hyperlipidemi* OR hyperlipidaemi*)[abs] OR (hypercholesterolemi* OR hypercholesterolaemi* OR hyperlipidemi* OR hyperlipidaemi*)[Keywords]) OR ((hyperlipoproteinemi* OR hyperlipoproteinaemi* OR hyperbetalipoproteinemi* OR hyperbetalipoproteinaemi*)[Title] OR (hyperlipoproteinemi* OR hyperlipoproteinaemi* OR hyperbetalipoproteinemi* OR hyperbetalipoproteinaemi*)[abs] OR (hyperlipoproteinemi* OR hyperlipoproteinaemi* OR hyperbetalipoproteinemi* OR

hyperbetalipoproteinaemi*)[Keywords]) OR ("Hyperlipidemias"[mh]) OR ("Hypercholesterolemia"[mh]) OR ("Hyperlipoproteinemia Type II"[mh]))) 11 hits

**Key:**
[abs] = abstract
[mh] = subject heading (MeSH heading)
[mhe] = exploded subject heading (MeSH heading)
* = truncation

**Science Citation Index**
via Web of Science, Clarivate Analytics https://clarivate.com/
Date range: 1900 - present
Date searched: 26th February 2026
Records retrieved: 1126

1: TS=(child or children or childhood* or infant* or infancy or pediatric* or paediatric* or preschool* or pre-school* or schoolchild* or school-age* or schoolage* or schoolboy* or schoolgirl*) Results: 2393783
2: TS=(girl or girls or boy or boys or kid or kids) Results: 236488
3: TS=(adolesc* or "young people" or "young person*" or teen* or youth* or preteen* or pubert* or prepubert* or pubescen* or prepubescen* or juvenil*) Results: 795770
4: TS=(neonat* or neo-nat* or newborn* or "new born*" or "newly born*" or baby or babies) Results: 544806
5: #1 OR #2 OR #3 OR #4 Results: 3249793
6: TS=((hyperlipoprotein$emi* or (hyper NEAR/3 lipoprotein$emi*) or hyperbetalipoprotein$emi*)) Results: 5360
7: TS=(hypercholesterol$emi* or hyper-cholesterol$emi*) Results: 56819
8: TS=(hyperlipid$emi* or hyper-lipid$emi*) Results: 44796
9: TS=heFH Results: 375
10: TS=("heterozygous FH") Results: 535
11: #6 OR #7 OR #8 OR #9 OR #10 Results: 99487
12: #11 AND #5 Results: 6227
13: TS=(statin*) Results: 72863
14: TS=(HMG-CoA or "hydroxymethylglutaryl CoA reductase inhibitor*" or "hydroxymethylglutaryl coenzyme a inhibitor*") Results: 14013
15: TS=(atorvastatin* or Lipitor* or lypqozet* or caduet*) Results: 20003
16: TS=(Fluvastatin* or lescol* or nandovar* or dorisin* or fluindostatin*) Results: 3030
17: TS=(lovastatin* or mevacor* or altoprev* or mevinolin*) Results: 8033
18: TS=(pitavastatin* or livalo* or zypitamag*) Results: 1713
19: TS=(pravastatin* or Pravachol*) Results: 9625
20: TS=(rosuvastatin* or crestor* or ezallor*) Results: 7659
21: TS=(simvastatin* or zocor* or flolipid* or vytorin* or inegy*) Results: 19402
22: #13 OR #14 OR #15 OR #16 OR #17 OR #18 OR #19 OR #20 OR #21 Results: 107862
23: #22 AND #12 Results: 840
24: TS=(ezetimibe* or ezetrol* or zetia*) Results: 6085
25: #24 AND #12 Results: 161
26: TS=((lipid-modif* or lipid-lower*) NEAR/2 (therap* or treatment* or intervention* or medication* or drug* or agent* or inject*)) Results: 17337
27: #26 AND #12 Results: 320
28: #27 OR #25 OR #23 Results: 1003

29: TS=((PCSK9* NEAR/3 (inhibitor* or block* or antagonist* or therap* or target*)) or PCSKi*) Results: 3671
30: TS=(("proprotein convertase subtilisin" or "kexin type 9") NEAR/3 (inhibitor* or block* or antagonist*)) Results: 1455
31: TS=("monoclonal antibod*" or mAb*) Results: 348379
32: TS=(Evolocumab* or repatha* or AMG-145 or AMG145 or sal-003 or sal003) Results: 1908
33: TS=(Alirocumab* or praluent* or SAR-236553 or SAR236553 or REGN727 or REGN-727) Results: 1393
34: TS=(enlicitide* or MK-0616 or MK0616) Results: 30
35: TS=(Inclisiran* or leqvio* or ALN-PCS* or ALNPCS* or aln-60212 or aln60212 or kjx-839 or kjx839) Results: 640
36: TS=((small or short) NEAR/2 ("interfering RNA" or "interfering ribonucleic acid")) Results: 34429
37: TS=(siRNA*) Results: 95608
38: TS=("bile acid sequest*") Results: 948
39: TS=(colesevelam* or cholestagel* or lodalis* or welchol or gt31-104* or gt-31-104* or iw-3718 or iw3718) Results: 536
40: TS=(colestilan* or bindren* or cholebine* or colestimide* or mci-196 or mci196) Results: 106
41: TS=(colestipol* or colestid* or cholestabyl* or lestid*) Results: 911
42: TS=(cholestyramin* or colestyramin* or cuemid* or quantalan* or questran* or olestyr* or prevalite* or mk-135 or mk135) Results: 3180
43: TS=(diethylaminoethyldextran* or diethylaminoethyl-dextran* or DEAE-dextran* or DEAE-sephadex* or pulsar* or dexide* or nolipid*) Results: 28831
44: TS=("bempedoic acid*" or bempedoate* or nexleto* or nilemdo* or nexlizet* or nustendi* or esp-55016 or esp55016 or etc-1002 or etc1002) Results: 741
45: TS=(lerodalcibep* or lerochol* or lib-003 or lib003) Results: 29
46: #45 OR #44 OR #43 OR #42 OR #41 OR #40 OR #39 OR #38 OR #37 OR #36 OR #35 OR #34 OR #33 OR #32 OR #31 OR #30 OR #29 Results: 497706
47: #46 AND #12 Results: 321
48: #47 OR #28 Results: 1144
49: TI=(animal or animals or rat or rats or mouse or mice or rodent or rodents or porcine or murine or sheep or lamb or lambs or ewe or ewes or pig or pigs or piglet or piglets or sow or sows or minipig or minipigs or rabbit or rabbits or kitten or kittens or dog or dogs or puppy or puppies or monkey or monkeys or horse or horses or foal or foals or equine or calf or calves or cattle or heifer or heifers or hamster or hamsters or chicken or chickens or livestock) Results: 3329669
50: #48 NOT #49 Results: 1126

**Key:**
TS = topic tag; searches in title, abstract, author keywords and keywords plus fields
TI = title search
* = truncation
$ = represents zero or one character
NEAR/3 = terms within three words of each other (any order)

***On-going, unpublished or grey literature search strategies***

**ClinicalTrials.gov**
https://clinicaltrials.gov/ct2/

Date searched: 26th February 2026
Records retrieved: 263

Advanced search page used with age group child (birth-17) box ticked.

263 Studies found for: hypercholesterolemia OR hypercholesterolaemia OR hyperlipoproteinemia OR hyperlipoproteinaemia OR hyperlipidemia OR hyperlipidaemia OR heFH OR "heterozygous FH"

**Conference Proceedings Citation Index – Science (CPCI-Science)**
via Web of Science, Clarivate Analytics https://clarivate.com/
Date range: 1990 - present
Date searched: 26th February 2026
Records retrieved: 65

1: TS=(child or children or childhood* or infant* or infancy or pediatric* or paediatric* or preschool* or pre-school* or schoolchild* or school-age* or schoolage* or schoolboy* or schoolgirl*) Results: 227073
2: TS=(girl or girls or boy or boys or kid or kids) Results: 12003
3: TS=(adolesc* or "young people" or "young person*" or teen* or youth* or preteen* or pubert* or prepubert* or pubescen* or prepubescen* or juvenil*) Results: 65225
4: TS=(neonat* or neo-nat* or newborn* or "new born*" or "newly born*" or baby or babies) Results: 37028
5: #1 OR #2 OR #3 OR #4 Results: 305756
6: TS=((hyperlipoprotein$emi* or (hyper NEAR/3 lipoprotein$emi*) or hyperbetalipoprotein$emi*)) Results: 193
7: TS=(hypercholesterol$emi* or hyper-cholesterol$emi*) Results: 5355
8: TS=(hyperlipid$emi* or hyper-lipid$emi*) Results: 3000
9: TS=heFH Results: 21
10: TS=("heterozygous FH") Results: 17
11: #6 OR #7 OR #8 OR #9 OR #10 Results: 8160
12: #11 AND #5 Results: 449
13: TS=(statin*) Results: 8981
14: TS=(HMG-CoA or "hydroxymethylglutaryl CoA reductase inhibitor*" or "hydroxymethylglutaryl coenzyme a inhibitor*") Results: 916
15: TS=(atorvastatin* or Lipitor* or lypqozet* or caduet*) Results: 2155
16: TS=(Fluvastatin* or lescol* or nandovar* or dorisin* or fluindostatin*) Results: 354
17: TS=(lovastatin* or mevacor* or altoprev* or mevinolin*) Results: 536
18: TS=(pitavastatin* or livalo* or zypitamag*) Results: 254
19: TS=(pravastatin* or Pravachol*) Results: 887
20: TS=(rosuvastatin* or crestor* or ezallor*) Results: 839
21: TS=(simvastatin* or zocor* or flolipid* or vytorin* or inegy*) Results: 1855
22: #13 OR #14 OR #15 OR #16 OR #17 OR #18 OR #19 OR #20 OR #21 Results: 14357
23: #22 AND #12 Results: 47
24: TS=(ezetimibe* or ezetrol* or zetia*) Results: 641
25: #24 AND #12 Results: 3
26: TS=((lipid-modif* or lipid-lower*) NEAR/2 (therap* or treatment* or intervention* or medication* or drug* or agent* or inject*)) Results: 1273
27: #26 AND #12 Results: 10
28: #27 OR #25 OR #23 Results: 56

29: TS=((PCSK9* NEAR/3 (inhibitor* or block* or antagonist* or therap* or target*)) or PCSKi*) Results: 310
30: TS=(("proprotein convertase subtilisin" or "kexin type 9") NEAR/3 (inhibitor* or block* or antagonist*)) Results: 52
31: TS=("monoclonal antibod*" or mAb*) Results: 19659
32: TS=(Evolocumab* or repatha* or AMG-145 or AMG145 or sal-003 or sal003) Results: 192
33: TS=(Alirocumab* or praluent* or SAR-236553 or SAR236553 or REGN727 or REGN-727) Results: 173
34: TS=(enlicitide* or MK-0616 or MK0616) Results: 2
35: TS=(Inclisiran* or leqvio* or ALN-PCS* or ALNPCS* or aln-60212 or aln60212 or kjx-839 or kjx839) Results: 95
36: TS=((small or short) NEAR/2 ("interfering RNA" or "interfering ribonucleic acid")) Results: 530
37: TS=(siRNA*) Results: 2740
38: TS=("bile acid sequest*") Results: 70
39: TS=(colesevelam* or cholestagel* or lodalis* or welchol or gt31-104* or gt-31-104* or iw-3718 or iw3718) Results: 75
40: TS=(colestilan* or bindren* or cholebine* or colestimide* or mci-196 or mci196) Results: 11
41: TS=(colestipol* or colestid* or cholestabyl* or lestid*) Results: 52
42: TS=(cholestyramin* or colestyramin* or cuemid* or quantalan* or questran* or olestyr* or prevalite* or mk-135 or mk135) Results: 136
43: TS=(diethylaminoethyldextran* or diethylaminoethyl-dextran* or DEAE-dextran* or DEAE-sephadex* or pulsar* or dexide* or nolipid*) Results: 5085
44: TS=("bempedoic acid*" or bempedoate* or nexleto* or nilemdo* or nexlizet* or nustendi* or esp-55016 or esp55016 or etc-1002 or etc1002) Results: 102
45: TS=(lerodalcibep* or lerochol* or lib-003 or lib003) Results: 13
46: #29 OR #30 OR #31 OR #32 OR #33 OR #34 OR #35 OR #36 OR #37 OR #38 OR #39 OR #40 OR #41 OR #42 OR #43 OR #44 OR #45 Results: 28846
47: #46 AND #12 Results: 12
48: #47 OR #28 Results: 65
49: TI=(animal or animals or rat or rats or mouse or mice or rodent or rodents or porcine or murine or sheep or lamb or lambs or ewe or ewes or pig or pigs or piglet or piglets or sow or sows or minipig or minipigs or rabbit or rabbits or kitten or kittens or dog or dogs or puppy or puppies or monkey or monkeys or horse or horses or foal or foals or equine or calf or calves or cattle or heifer or heifers or hamster or hamsters or chicken or chickens or livestock) Results: 309706
50: #48 NOT #49 Results: 65

**Key:**
TS = topic tag; searches in title, abstract, author keywords and keywords plus fields
TI = title search
* = truncation
$ = represents zero or one character
NEAR/3 = terms within three words of each other (any order)

**EU Clinical Trials Register**
https://www.clinicaltrialsregister.eu/ctr-search/search
Search date: 26th February 2026
Records retrieved: 46

Advanced search with following age categories selected – adolescent, children, infant and toddler, newborn, preterm newborn infants, under 18.

1. 46 result(s) found for: (hypercholesterolemia OR hypercholesterolaemia OR hyperlipoproteinemia OR hyperlipoproteinaemia OR hyperlipidemia OR hyperlipidaemia OR heFH OR "heterozygous FH")

**PROSPERO**
via https://www.crd.york.ac.uk/prospero/
Date searched: 26th February 2026
Records retrieved: 167

filtered to clinical not animal = 167
17 ongoing
150 completed

73 #72 or #44 168
72 #71 and #17 85
71 #70 or #69 or #68 or #67 or #66 or #65 or #64 or #63 or #62 or #61 or #60 or #59 or #58 or #57 or #56 or #55 or #54 or #53 or #52 or #51 or #50 or #49 or #48 or #47 or #46 or #45 4345
70 lerodalcibep* or lerochol* or lib-003 or lib003 8
69 "bempedoic acid" or bempedoate* or nexleto* or nilemdo* or nexlizet* or nustendi* 94
68 diethylaminoethyldextran* or diethylaminoethyl-dextran* or DEAE-dextran* or DEAE-sephadex* or pulsar* or dexide* or nolipid* 14
67 MeSH DESCRIPTOR DEAE-Dextran 0
66 cholestyramin* or colestyramin* or cuemid* or quantalan* or questran* or olestyr* or prevalite* 39
65 MeSH DESCRIPTOR Cholestyramine Resin 3
64 colestipol* or colestid* or cholestabyl* or lestid* 20
63 MeSH DESCRIPTOR Colestipol 1
62 colestilan* or bindren* or cholebine* or colestimide* 6
61 colesevelam* or cholestagel* or lodalis* or welchol* 14
60 MeSH DESCRIPTOR Colesevelam Hydrochloride 2
59 "bile acid" adj sequest* 68
58 siRNA* 170
57 (small or short) adj2 ("interfering RNA" or "interfering ribonucleic acid") 84
56 MeSH DESCRIPTOR RNA, Small Interfering 54
55 Inclisiran* or leqvio* or ALN-PCS* or ALNPCS* or aln-60212 or aln60212 or kjx-839 or kjx839 155
54 enlicitide* or MK-0616 or MK0616 13
53 Alirocumab* or praluent* or SAR-236553 or SAR236553 or REGN727 or REGN-727 193
52 Evolocumab* or repatha* or AMG-145 or AMG145 or sal-003 or sal003 190
51 (monoclonal adj antibod*) or mAb* 3052
50 MeSH DESCRIPTOR Antibodies, Monoclonal, Humanized 703
49 MeSH DESCRIPTOR Antibodies, Monoclonal 803
48 "kexin type 9" adj3 (inhibitor* or block* or antagonist*) 117
47 "proprotein convertase subtilisin" adj3 (inhibitor* or block* or antagonist*) 6
46 PCSKi* 1
45 PCSK9* adj3 (inhibitor* or block* or antagonist* or therap* or target*) 416
44 #43 or #39 or #35 160
43 #42 and #17 111
42 #40 or #41 959

41 lipid-lower* adj2 (therap* or treatment* or intervention* or medication* or drug* or agent* or inject*) 926
40 lipid-modif* adj2 (therap* or treatment* or intervention* or medication* or drug* or agent* or inject*) 57
39 #38 and #17 52
38 #37 or #36 379
37 ezetimibe* or ezetrol* or zetia*379
36 MeSH DESCRIPTOR Ezetimibe EXPLODE 115
35 #34 and #17 134
34 #18 or #19 or #20 or #21 or #22 or #23 or #24 or #25 or #26 or #27 or #28 or #29 or #30 or #31 or #32 or #33 3352
33 statin* 3170
32 simvastatin* or zocor* or flolipid* or vytorin* or inegy* 340
31 rosuvastatin* or crestor* or ezallor* 311
30 pravastatin* or Pravachol* 216
29 pitavastatin* or livalo* or zypitamag* 138
28 lovastatin* or mevacor* or altoprev* or mevinolin* 176
27 Fluvastatin* or lescol* or nandovar* or dorisin* or fluindostatin* 171
26 atorvastatin* or Lipitor* or lypqozet* or caduet* 430
25 HMG-CoA or ("hydroxymethylglutaryl CoA reductase" adj inhibitor*) or ("hydroxymethylglutaryl coenzyme a" adj inhibitor*) 940
24 MeSH DESCRIPTOR Simvastatin EXPLODE 61
23 MeSH DESCRIPTOR Rosuvastatin Calcium 53
22 MeSH DESCRIPTOR Pravastatin 34
21 MeSH DESCRIPTOR Lovastatin 21
20 MeSH DESCRIPTOR Fluvastatin 15
19 MeSH DESCRIPTOR Atorvastatin 93
18 MeSH DESCRIPTOR Hydroxymethylglutaryl-CoA Reductase Inhibitors EXPLODE 911
17 #16 and #8 638
16 #15 or #14 or #13 or #12 or #11 or #10 or #9 133953
15 neonat* or neo-nat* or newborn* or "new born" or "new borns" or "newly born" or baby or babies 18308
14 adolesc* or "young people" or "young person" or "young persons" or teen* or youth* or preteen* or pubert* or prepubert* or pubescen* or prepubescen* or juvenil* 53656
13 girl or girls or boy or boys or kid or kids 3581
12 child or children or childhood* or infant* or infancy or pediatric* or paediatric* or preschool* or pre-school* or schoolchild* or school-age* or schoolage* or schoolboy* or schoolgirl* 111504
11 MeSH DESCRIPTOR Infant EXPLODE 7993
10 MeSH DESCRIPTOR Adolescent 13877
9 MeSH DESCRIPTOR Child EXPLODE 26370
8 #1 or #2 or #3 or #4 or #5 or #6 or #7 2539
7 heFH or "heterozygous FH" 59
6 hyperlipid*emi* or hyper-lipid*emi* 1800
5 hypercholesterol*emi* or hyper-cholesterol*emi* 986
4 (hyperlipoprotein*emi* or (hyper adj3 lipoprotein*emi*) or hyperbetalipoprotein*emi*) 120
3 MeSH DESCRIPTOR Hypercholesterolemia 160
2 MeSH DESCRIPTOR Hyperlipidemias 338
1 MeSH DESCRIPTOR Hyperlipoproteinemia Type II 76

**Key:**
MeSH DESCRIPTOR = subject heading (MeSH heading)
* = truncation
adj3 = terms within 3 words of each other (order specified)

**WHO International Clinical Trials Registry Platform (ICTRP)**
https://trialsearch.who.int/AdvSearch.aspx
Date searched: 26th February 2026
Records retrieved: 205

Advanced search screen. Recruitment status set to ALL, restricted to clinical trials in children, search without synonyms.

1.Condition field: hypercholesterolemia* OR hypercholesterolaemia* OR hyperlipoproteinemia* OR hyperlipoproteinaemia* OR hyperlipidemia* OR hyperlipidaemia* OR heFH OR "heterozygous FH"
205 trials

***Guideline website searches***

Simple searches were carried out via the guideline websites listed below and any results were browsed for relevance. Relevant guidelines identified were checked against the EndNote library of results and added to the library if they had not already been found through previous searches.

**National Institute of health and Care Excellence (NICE)**
https://www.nice.org.uk/
Date searched: 26th February 2026
Records retrieved: 8

1. "Familial hypercholesterolaemia", limited to guidance documents – 13 results browsed for relevance, 8 relevant.

**Trip database**
https://www.tripdatabase.com/
Date searched: 26th February 2026
Records retrieved: 47

hypercholesterolaemia OR hypercholstolerolemia OR hefh OR "heterozygous fh" OR hyperlipoproteinemia OR hyperlipoproteinaemia OR hyperlipidemia OR hyperlipidaemia

**GIN international guideline library**
https://guidelines.ebmportal.com/
Date searched: 26th February 2026
Records retrieved: 8

1. "familial hypercholesterolemia" – 0 hits
2. "familial hypercholesterolaemia" – 0 hits
3. hypercholesterolemia – 1 hit
4. hypercholesterolaemia – 2 hits, 1 was a duplicate
5. heterozygous FH – 0 hits

6. HeFH - 0 hits
7. Hyperlipoproteinemia – 0 hits
8. Hyperlipoproteinaemia - 0 hits
9. hyperlipidemia – 4 results, 2 were duplicates
10. hyperlipidaemia – 0 hits
11. dyslipidaemia – 3 hits, 2 were duplicates
12. dyslipidemia – 6 hits, 4 were duplicates
